# The use of residual blood specimens in seroprevalence studies for vaccine preventable diseases: A scoping review

**DOI:** 10.1101/2025.02.10.25321868

**Authors:** Monica Pilewskie, Christine Prosperi, Abigail Bernasconi, Ignacio Esteban, Lori Niehaus, Connor Ross, Andrea C Carcelén, William J Moss, Amy K Winter

## Abstract

**Background:** Residual blood specimens offer a cost- and time-efficient alternative for conducting serological surveys. However, their use is often criticized due to potential issues with representativeness of the target population and/or limited availability of associated metadata. We conducted a scoping review to examine where, when, how, and why residual blood specimens have been used in serological surveys for vaccine-preventable diseases (VPDs), and how potential selection biases are addressed.

**Methods:** The review followed the Preferred Reporting Items for Systematic Reviews and Meta-Analyses extension for Scoping Reviews (PRISMA-ScR). We identified relevant papers published between 1999 and 2022 through a literature search of PubMed, Scopus, Embase, Cochrane, and the WHO IRIS database. Study data were captured using Kobo Toolbox, and findings were summarized using descriptive analytical methods.

**Results:** A total of 601 articles met the inclusion criteria after title, abstract screening, and full-text review. The most commonly studied VPDs using residual blood specimens were COVID-19 (27%), hepatitis E (16%), hepatitis B (10%), influenza (9%), HPV (7%), and measles (7%). Most studies (81%) aimed to estimate population-level seroprevalence. Residual specimens were primarily sourced from patients (55%) or blood donors (36%). Common strategies to address potential biases included comparing results with published estimates (78%) and performing stratified analyses (71%).

**Conclusions:** Residual blood specimens are widely used in seroprevalence studies, particularly during emerging disease outbreaks when rapid estimates are critical. However, the review highlighted inconsistencies in how researchers analyze and report the use of residual specimens. To address these gaps, we propose a set of recommendations to improve the analysis, reporting, and ethical considerations of serological surveys using residual specimens.

## Introduction

Immunoglobulin G (IgG) antibodies can be used to identify persons with prior exposure to vaccine preventable diseases (VPDs) through infection or vaccination. Serological surveys (serosurveys) link serological testing of IgG antibodies results with individual demographic and epidemiologic data. These rich data sets can be used to directly estimate population seroprevalence profiles by characteristics of interest (e.g., age or space) and identify immunity gaps. Serosurveys can also be used to estimate disease burden [1,2], infer key epidemiological parameters (e.g., basic reproduction number or rate of infection) [1,3,4], evaluate surveillance and vaccination program performance [5,6], and estimate outbreak risk [7,8]. The results generated from high-quality serosurveys of VPDs can inform vaccination strategies and control the spread of disease [9].

High-quality serosurveys are population-based, household surveys which use probability-based sampling designs to accurately include a representative sample of the population of interest. They require standardized laboratory methods with excellent quality assurance and control for the data to be appropriately analyzed and interpreted. The main limitations to conducting high-quality serosurveys include the substantial staff and resource requirements; the necessary sampling, laboratory, and analytic capacity; and the long timeframe needed to plan, conduct, test, analyse, and disseminate the results [10,11]. Consequently, routinely collected vaccination coverage and case surveillance data are relied upon to inform vaccination programs. While immunity profiles can be indirectly estimated for vaccine preventable diseases given historical vaccination coverage and surveillance data, the quality of these data are highly variable [12], and serosurveys provide a less biased and more direct estimation of immunity profiles [13,7].

One way to increase the feasibility of serosurveys is to use residual blood specimens. Residual blood consists of remnant specimens collected for another purpose (for example, blood donations, past serosurveys, or other laboratory tests) that are available for an objective not originally intended. Using residual specimens in serosurveys reduces the resource commitment, including time and staff for specimen collection, which is often the highest cost of a serosurvey [11]. Residual specimens also protect data collectors from exposure to emergent pathogens, a concern during the COVID-19 pandemic [14,15]. However, a critical limitation of using residual specimens is the potential for a biased sample that is not representative of the population of interest and conclusions that lack external validity.

In this scoping review, we evaluated the use of residual specimens in seroprevalence studies of VPDs. Specifically, we aimed to 1) characterize serological studies that used residual specimens; 2) describe the objectives of serological studies that use residual specimens; and 3) evaluate if and how authors handled or discussed potential selection bias. This review follows the authors’ observations that since the emergence of COVID-19, there has been an increase in published papers on serosurveys that use residual specimens. Additionally, with the development of multiplex technology for testing serum for antibodies to multiple antigens and integrated sero-surveillance systems [16], there is potential for residual specimens to be more widely used to understand a broad array of pathogen exposures to guide interventions to better control the spread of infectious diseases. We sought to systematically evaluate our observation of an increase in serosurveys using residual specimens and understand the past and potential future role of residual specimens in the field of sero-epidemiology to inform surveillance and response to outbreaks of VPDs.

## Materials and Methods

We summarize the protocol below but see Text S1 for the full protocol. This review is reported according to the Preferred Reporting Items for Systematic Reviews and Meta-Analyses extension for Scoping Reviews (PRISMA-ScR) statement (Text S2).

### Inclusion and exclusion criteria

The inclusion criteria were studies that used residual human blood for serological testing of pathogens causing VPDs of interest: *Vibrio cholerae* (cholera), dengue viruses, *Corynebacterium diphtheriae* (diphtheria), hepatitis A virus, hepatitis B virus, hepatitis E virus, *Haemophilus influenza* type b (Hib), human papillomavirus virus (HPV), influenza virus, Japanese encephalitis virus, measles virus, *Neisseria meningitidis* (meningococcal meningitis), mumps virus, *Bordetella pertussis* (pertussis), *Streptococcus pneumoniae* (pneumococcus), poliovirus (poliomyelitis), rotavirus, rubella virus, SARS-CoV-2 virus (COVID-19), *Clostridium tetani* (tetanus), tick-borne encephalitis virus, *Mycobacterium tuberculosis* (tuberculosis), *Salmonella typhi* (typhoid fever), varicella-zoster virus (herpes zoster/shingles & varicella/chickenpox), and yellow fever virus. We defined serosurveys using residual blood as studies that tested remnant, surplus, existing, archived, or left-over blood samples for purposes beyond the original reason for sample collection. The serological testing must have been conducted to measure exposure or immune status and not acute infection; for example, testing conducted with IgG immunoassays, neutralization, or hemagglutinin-inhibition tests and not nucleic acid detection assays or IgM immunoassays. The objective of this scoping review was to evaluate the use of residual blood in serological surveys measuring population-level metrics; therefore, we excluded studies that used residual blood to evaluate diagnostic tests [17] or inform diagnostic test laboratory protocols, or that used residual blood as a comparison group [18,19].

### Information sources and literature search

The screening of articles was conducted using English, French, Spanish, Italian, Russian, Chinese, and Portuguese language literature, including peer-reviewed literature, grey literature, and pre-prints, between January 1990 and August 2022. We excluded editorials, letters, commentaries, narrative reviews, and conference abstracts. Initial screening was conducted in June 2021 and re-run in August 2022 to include studies published after the initial screening. Using search terms for vaccine-preventable diseases, serologic testing, and residual samples, the following electronic databases were used: PubMed, Scopus, Embase, Cochrane, and the World Health Organization (WHO) Institutional Repository for Information Sharing (IRIS) database. Additional studies were identified through suggestions from experts at the United States Centers for Disease Control and Prevention (CDC) and WHO. We worked with librarians at Johns Hopkins University to create a list of search terms based on our inclusion and exclusion criteria. Database-specific search terms can be found in the protocol (see Text S1).

### Screening process

The screening criteria were established per the protocol and standardized among the investigators. After deleting duplicates, two members of the literature review group systematically screened the title and abstract of papers for the inclusion and exclusion criteria using Covidence systematic review software [20]. Those that met the criteria underwent full text review. The title and abstract and full-text screenings were done by two reviewers with disagreements resolved through discussion and arbitration with a third reviewer.

### Data extraction process and data points

A data extraction form was developed *a priori* utilizing KoboToolbox data collection platform and later calibrated following full text review (see Text S3 for data extraction survey). Data were extracted by one reviewer. A second reviewer was consulted if questions arose about adherence to inclusion and exclusion criteria or the data extraction process. Main data points collected included: VPD of interest, specimen countries of origin, year of original sample collection, the objectives of the serological study based on the abstract or summary alone (see Table S2 for more detail), original population from which the specimens were collected (see Table S1 for more detail), meta-data linked to residual samples (e.g. age, sex), permission and ethical considerations around testing residual samples, the investigator’s approach to exploring, discussing, or addressing potential selection biases (see Table S3 for more detail), and if collection was part of a larger serological surveillance system (see Text S4). No attempts were made to contact study investigators to obtain or confirm data or critically appraise the source of data.

### Data synthesis

Data synthesis was conducted using descriptive statistics. Data cleaning was conducted with SAS Version 9.4. All figures were created with the R statistical computing software (version 4.3.1; R Core Team 2023) using packages *ggplot2* (version 1.1.2), *sf* (version 1.0.14), *rnaturalearthdata* for the global shapefile (version 0.1.0), and *unikn* (version 0.8.0) to define color palettes.

## Results

The initial and secondary database searches yielded 11,526 unique articles eligible for screening (Figure 1). Of the 11,526 articles, 10,165 (88%) were excluded via title and abstract review. Among the 1,360 articles in full text review, 326 (24%) were excluded because the study did not use residual specimens, 302 (22%) were editorials, commentaries, or narrative reviews, and 114 articles (8%) were excluded based on other remaining criteria. Data extraction was conducted on 601 unique articles to be included in the final analysis (see Table S4 for a list and bibliography of all 601 articles).

**Figure 1.**
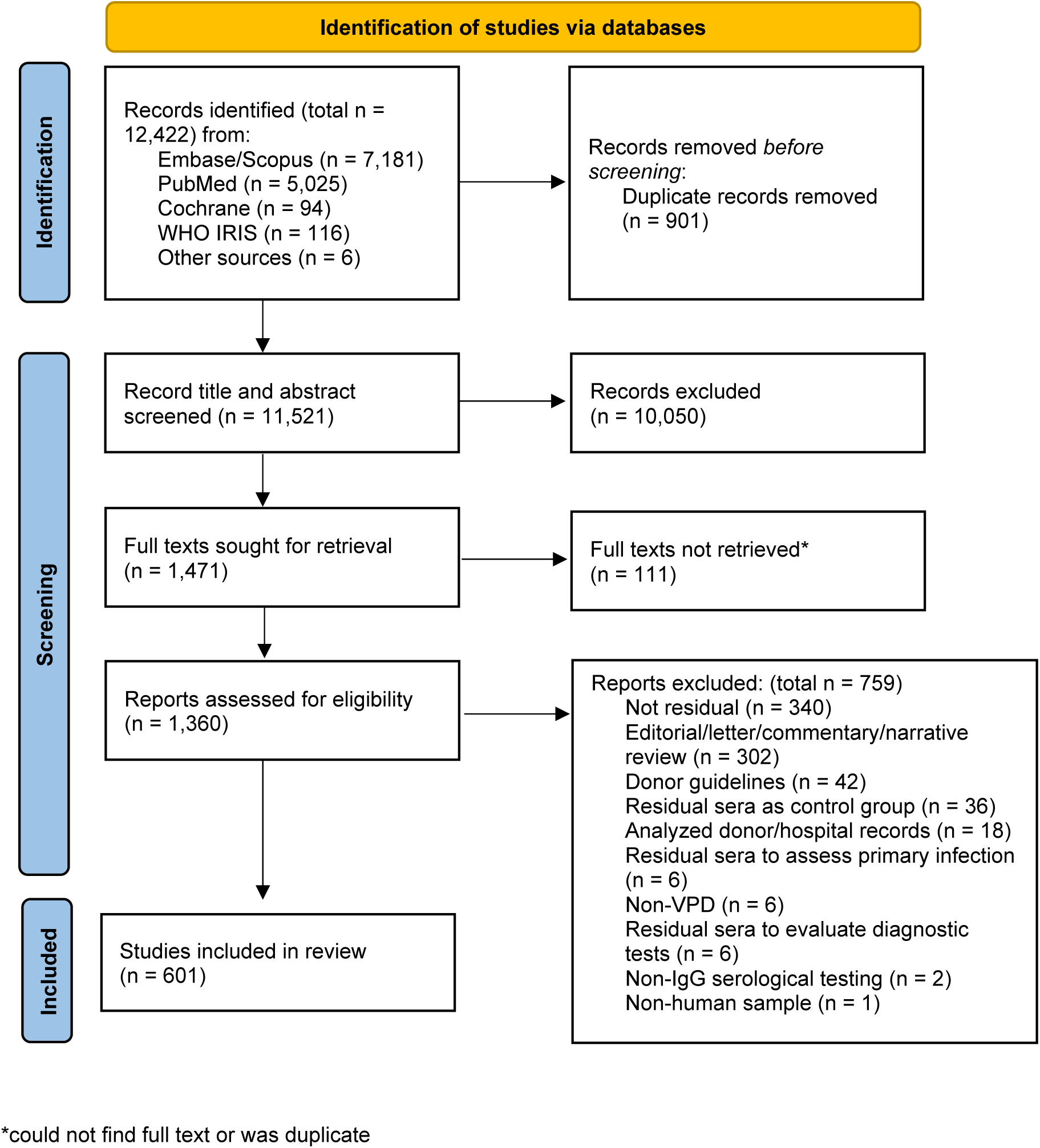
Flow Diagram of seroprevalence studies in the scoping review. *From:* Page MJ, McKenzie JE, Bossuyt PM, Boutron I, Hoffmann TC, Mulrow CD, et al. The PRISMA 2020 statement: an updated guideline for reporting systematic reviews. BMJ 2021;372:n71. doi: 10.1136/bmj.n71

### VPDs studied, trends, and objectives

Over 75% of the articles studied one or more of the following six VPDs: COVID-19, hepatitis E, hepatitis B, influenza, HPV, and measles. COVID-19 was the most common vaccine preventable disease studied between 1990 and 2022 (27.1% of articles), even though articles on COVID-19 were only published beginning in 2020 (Table 1). Seroprevalence studies of hepatitis E, hepatitis B, influenza, HPV, and measles were the next most common VPDs using residual specimens (Table 1). Overtime, an increase was observed in the number of published serological studies that use residual specimens (Figure 2A). The increase in serological articles using residual blood after 2020 was due to serological studies on COVID-19 (Figure 2B). There was an increase in articles studying measles from 2019 to 2020 during the global resurgence, hepatitis E from 2018 to 2019 following outbreaks in Bangladesh and South Sudan, and influenza from 2010 to 2011 following the 2009 H1N1 influenza virus pandemic (Figure 2B). The objective of serological surveys that used residual specimens was most often to describe population seroprevalence (80.5%), followed by the desire to identify risk factors for seropositivity (33.3%), estimate infection rates (17.6%), and evaluate trends in seropositivity (15.8%) (Figure 3, Text S5, Table S5). Studies on emergent pathogens such as COVID-19 and influenza disproportionally focused on the latter two objectives.

**Figure 2.**
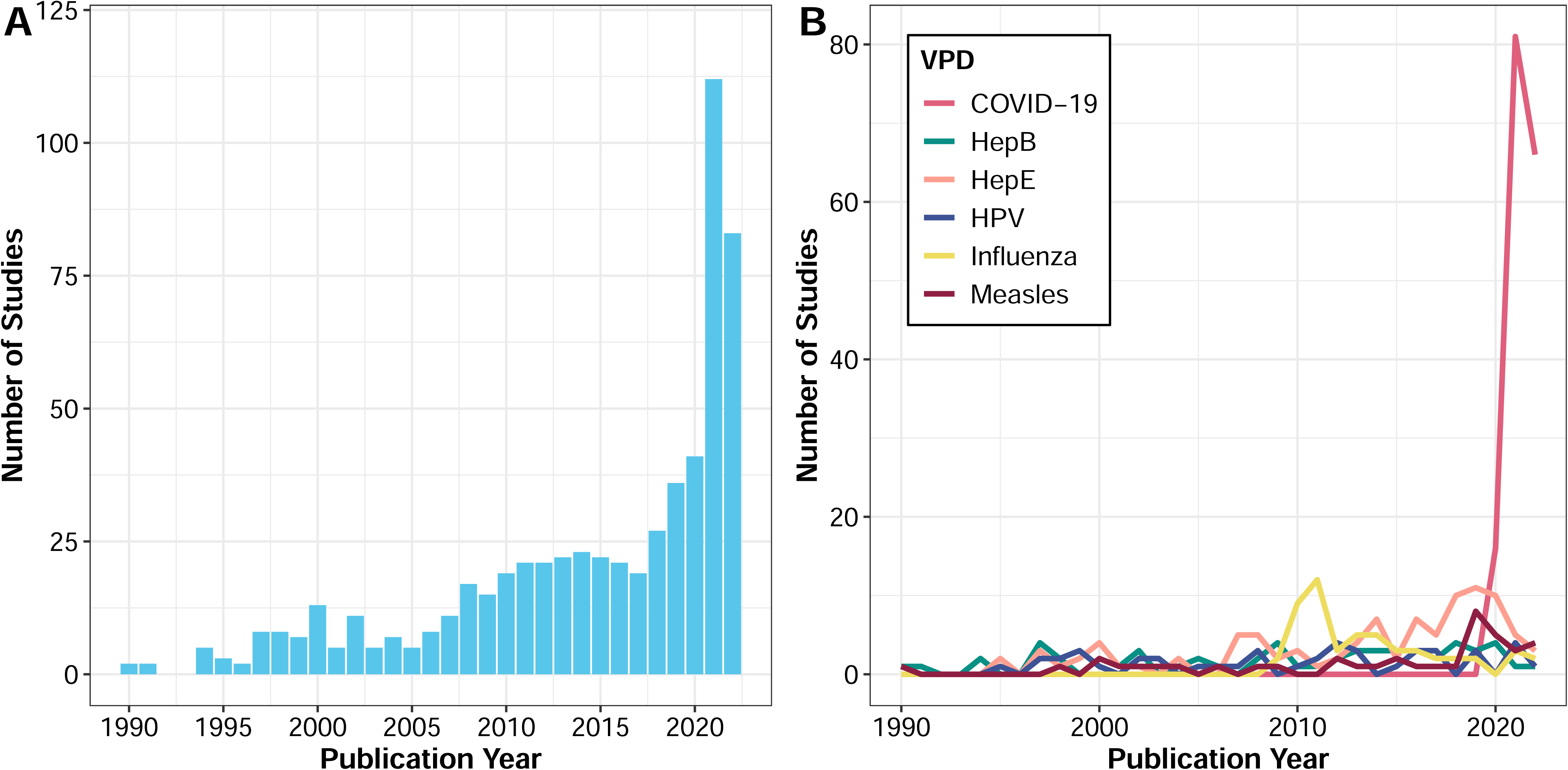
Time series of studies’ publication year A) across all VPDs (N = 601 studies) and B) by the six most studied VPDs.

**Figure 3.**
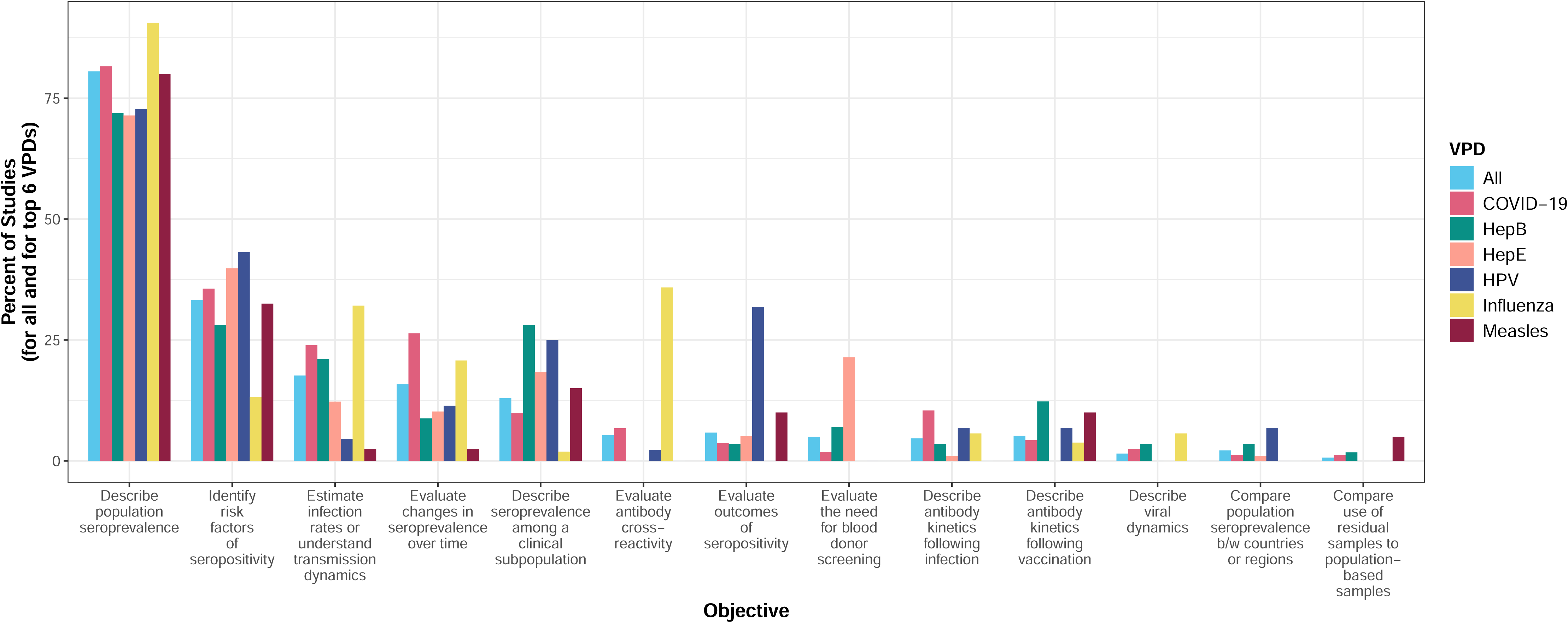
Bar plot displaying percent of studies by objective for all VPDs and for the six most studied VPDs (N = 601 studies). Note the categories are not mutually exclusive meaning that one study could have conducted testing for multiple pathogens and have had multiple objectives. Refer to Table S2 for additional information.

**Table 1.** Number of articles reporting serological surveys using residual blood specimens by VPD. Note the categories below are not mutually exclusive meaning that one article could have conducted testing for multiple pathogens.

| **Vaccine-Preventable Disease** | **Total no. (%)**<br>**(N = 601)** |
| --- | --- |
| COVID-19 | 163 (27.1) |
| Hepatitis E (HepE) | 98 (16.3) |
| Hepatitis B (HepB) | 57 (9.5) |
| Influenza | 53 (8.8) |
| Human papillomavirus (HPV) | 44 (7.3) |
| Measles | 40 (6.7) |
| Hepatitis A (HepA) | 39 (6.5) |
| Rubella | 33 (5.5) |
| Dengue | 31 (5.2) |
| Varicella/Herpes zoster | 30 (5) |
| Diphtheria | 19 (3.2) |
| Mumps | 17 (2.8) |
| Pertussis | 17 (2.8) |
| Poliomyelitis | 8 (1.3) |
| Tetanus | 7 (1.2) |
| Japanese encephalitis | 4 (0.7) |
| Hemophilus influenza type b (Hib) | 3 (0.5) |
| Yellow fever | 3 (0.5) |
| Meningococcal meningitis | 2 (0.3) |
| Tick-borne encephalitis | 2 (0.3) |
| Rotavirus | 1 (0.2) |
| Typhoid fever | 1 (0.2) |
| Cholera | 0 (0.0) |
| Tuberculosis | 0 (0.0) |
| Pneumococcal | 0 (0.0) |
| Studies investigating single pathogen | 560 (93.2) |
| Studies investigating multiple pathogens | 41 (6.8) |

### Original use of specimens

The original use of residual specimens for most articles was as clinical diagnostic specimens (Figure 4). However, the most common original use for hepatitis E articles was for blood and plasma donations. Sixteen percent of specimens were originally collected for research purposes (serological survey or non-serological survey). This included surveys for specific medical conditions (e.g., hypertension, heart disease, lead exposure) or VPDs besides what the residual specimens were subsequently tested for.

**Figure 4.**
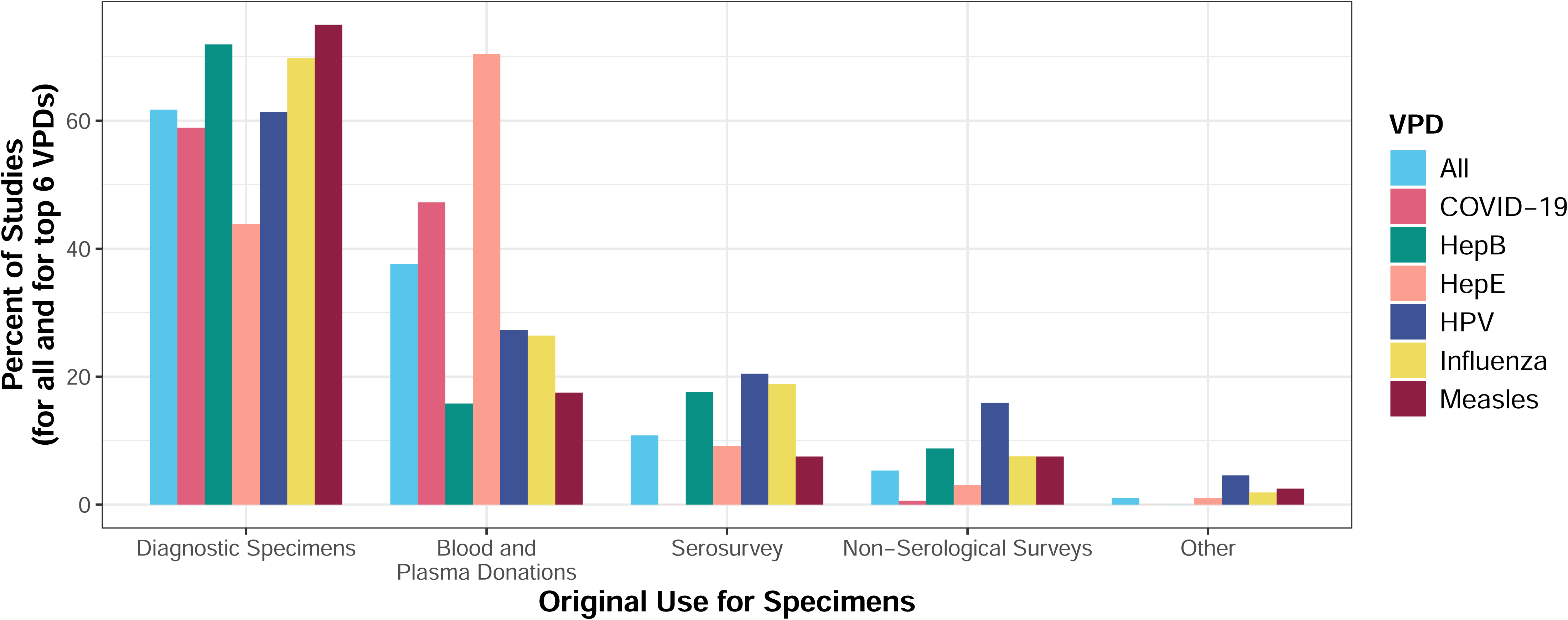
Bar plot displaying percent of studies by original use of specimens for all VPDs and for the six most studied VPDs (N = 601 studies). Note the categories are not mutually exclusive meaning that one study could have conducted testing for multiple pathogens or have use specimens from multiple original sources.

### Sources of residual specimens

The original population from which specimens were collected, and the collection site, was highly related to the original purpose for specimen collection. For example, given that most of the residual specimens’ original use was for diagnostic specimens, the main sources of residual specimens were patient populations from hospitals, clinics or diagnostic centers (see Text S5, Figures S1-S2, Table S6). The mean time between collection of specimens and publication of the study was 6.5 years but the time between collection and publication was faster for COVID-19 at 1.7 years (Text S5, Figure S3). In terms of location of source, a higher proportion of studies were conducted in the United States than in any other country, making up 14% (85 of 601) of articles published between 1990 and 2022 (Figure 5). Studies using specimens collected in low- and middle-income countries made up 35% (210 of 601) of articles. Twenty-eight (4.7%) of the 601 articles reported the testing was part of a larger serological surveillance system (Text S4, Table S7), including the Health Protection Agency National Seroepidemiology Program from the UK (12 articles) European Seroepidemiology Network (ESEN) (11 articles), Australia’s National Centre for Immunisation Research and Surveillance (4 articles) and Vietnam’s serosurveillance (1 article).

**Figure 5.**
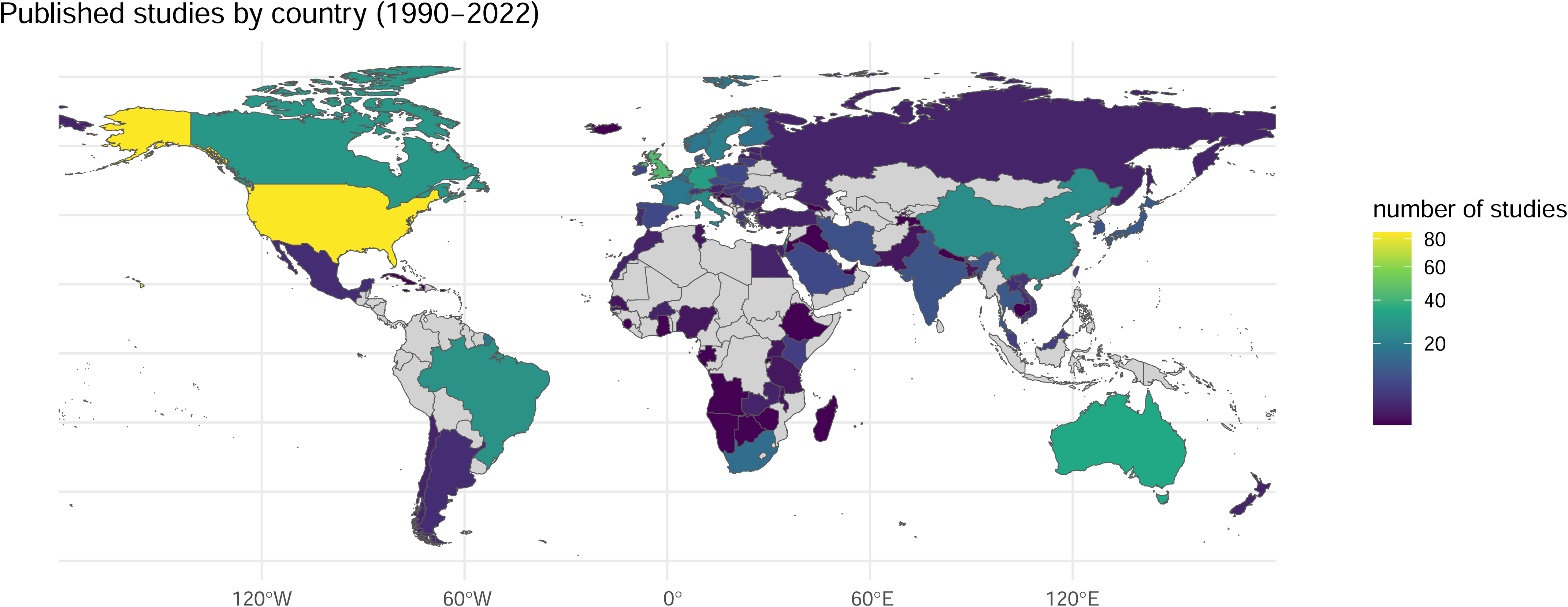
Map of published studies by country of original source population (N = 601 studies).

### Meta-data linked to residual specimens

Almost all articles used specimens linked to basic demographic data (e.g., age and/or sex), with 47% having access to extended demographic data such as geographic location (Table S8). When specimens from a previous serosurvey were used (11% of articles), 55% and 42% of these articles had access to extended demographic and epidemiologic meta-data (e.g., vaccination status), respectively. However, most articles (92%) used diagnostic specimens or specimens from blood or plasma donations and fewer than half (46%) had extended meta-data and only 11% had epidemiologic data.

### Ethical considerations in the use of residual specimens

Just over two-thirds (70.4%) of the articles reported receiving approval by an ethics board or committee to access and test the residual specimens (Table S9), with 35% of those studies using specimens from the United States, Australia, Canada, or the United Kingdom (results not shown). One-third (33.1%) of articles mentioned that broad individual consent for additional testing was obtained at the time of specimen collection, with most collected for blood or plasma donation or were clinical specimens; only 21.6% were from a survey. One-third (33.1%) mentioned the residual specimens were deidentified; among these all reported at least one additional ethical step was taken (72% indicating a committee approved additional testing, 27% indicated broad individual consent for additional testing, 14% received waiver for reconsenting, and 4% were exempt as public health surveillance). Studies that used specimens collected for clinical purposes were least likely to report that broad individual consent for additional testing was obtained but were most likely to report specimens were deidentified. Fifteen percent of articles did not include any statement on ethics or permissions.

### Selection Bias

A variety of approaches were used to explore or address potential selection bias due to the use of residual specimens (Table S3). Figure 5 shows the percentage of studies that use different approaches to address or handle bias overall and by the top six VPDs. The most common approach was to compare results to other published estimates (77.5%) (Figure 6). Conducting stratified analyses to control for differences in the sample and target populations (e.g., by age, sex, or location) was the second most common method (71%) across all VPD groups. About one quarter of studies used design-related considerations, such as stratified subsampling, to better reflect the population of interest (24.6%); this approach was particularly prevalent for influenza and measles studies. For example, Carcélen et al 2022 sub-sampled HIV serosurvey specimens in Zambia by age, province, and HIV infection status to obtain a representative sample and estimate measles seroprevalence by age and province [8]. Another frequently used approach included weighting results using characteristics linked to residual specimens (e.g., age, sex, or location) to align with the distribution of characteristics in the target population (22%). For example, Ho et al. 2020 estimated seroprevalence to hepatitis E virus in Belgium through weighting by province and age to account for differences in sampling [21]. Some studies (12.8%) relied on inclusion and exclusion criteria to reduce selection bias from the use of residual specimens (e.g., exclusion of specimens from a specific medical ward at a facility, specimens from patients admitted for respiratory illness, or patients who were immunocompromised [22]). Sensitivity analyses that varied the input data, model assumptions, or model parameters, were used in only 1% of the studies. For example, a study that examined associations between esophageal squamous cell carcinoma and HPV serological markers in serum from six existing case–control studies, conducted sensitivity analysis by excluding two studies in which blood collection procedures differed for the case and control subjects [23]. Lastly, some studies used a non-biased sample of specimens originating from the population of interest, particularly among hepatitis E and HPV studies. For example, a hepatitis E serosurvey used residual blood donor specimens to evaluate the need to include hepatitis E in blood donor screening [24], and an HPV serosurvey used residual diagnostic specimens from patients with confirmed squamous cell carcinoma of the oropharynx to assess the risk of oropharyngeal cancer as an outcome of HPV seropositivity [25].

**Figure 6.**
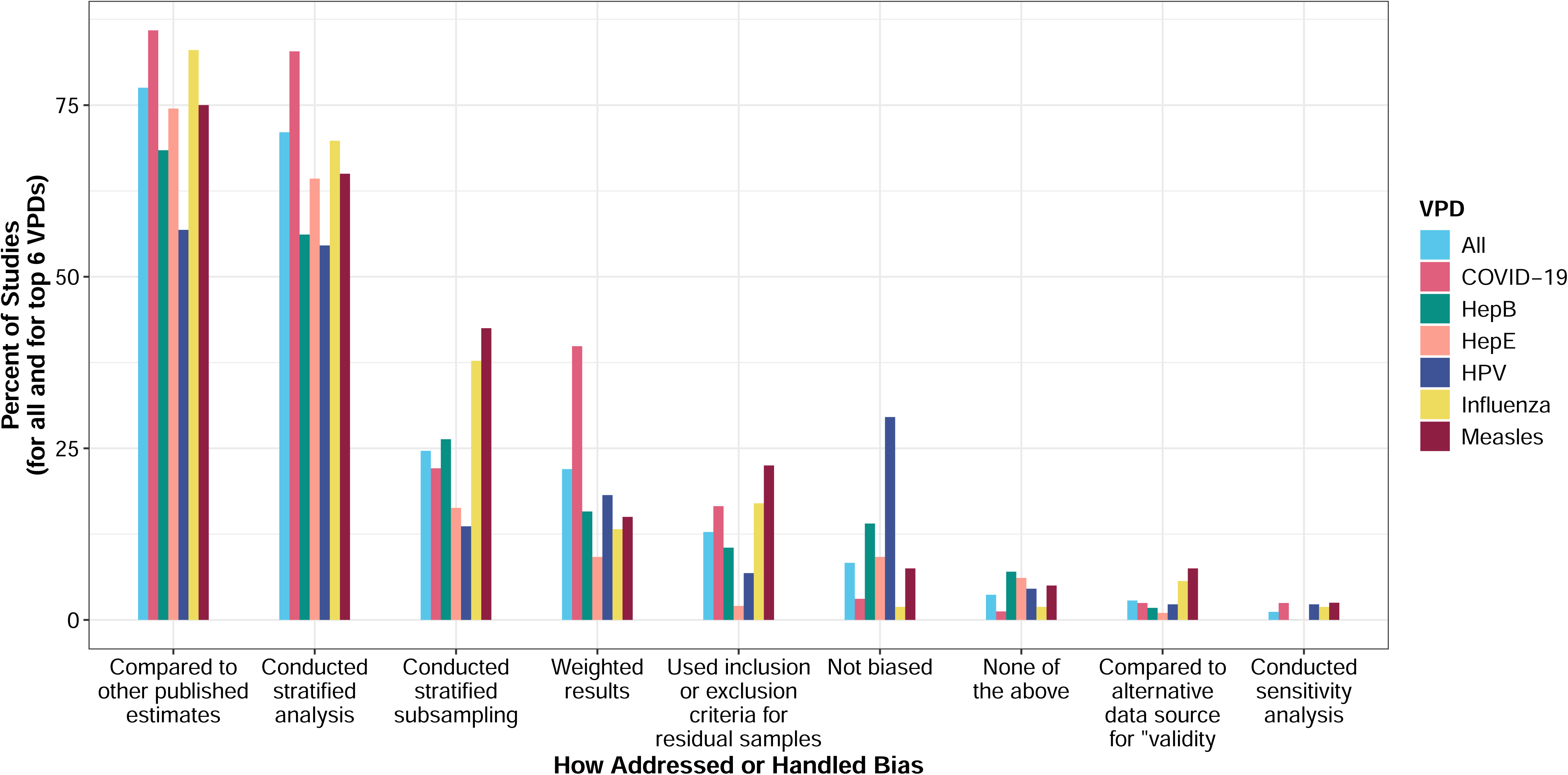
Exploring or addressing selection bias for all VPDs and for the six most studied VPDs (N = 601 studies). Note the categories are not mutually exclusive meaning that one study could have conducted testing for multiple pathogens or have discussed or handled bias in multiple ways. Refer to Table S3 for additional information.

## Discussion

We sought to characterize serological studies of VPDs that use residual blood specimens. The benefits of high-quality population-based serological surveys to understand and inform the control of infectious diseases are well documented [9,26]. However, high quality population-based serosurveys require resources of time, money, technical expertise, and cooperation of the population [11,27,28]. Residual specimens can be leveraged to reduce the time and cost of serosurveys but the potential lack of representativeness of the population of interest and limited meta-data are limitations.

Over 75% of studies addressed one or more of the following six VPDs: COVID-19, hepatitis E, and hepatitis B, influenza, HPV, and measles. Many studies were conducted during the 2009-2010 H1N1 influenza and 2019-2021 COVID-19 pandemics. The number of measles serosurveys using residual specimens increased in 2019 and 2020 during a global increase in the number of measles outbreaks. For example, Bassal et al. 2021 used stored samples from 2015 to assess measles seropositivity among an Israeli population to understand age-specific incidence rates of the 2018–2019 measles outbreak [29]. This increase in the number of serosurveys using residual specimens for COVID-19, influenza, and measles highlights the use of residual specimens to provide rapid information during emergent or re-emergent infectious disease outbreaks. This is particularly true when new data collection may be risky or difficult to conduct. For example, Uyoga et al. 2021 used residual specimens from Kenyan blood donations April-June 2020 to estimate COVID-19 seroprevalence and burden of disease during a time of movement restrictions in the country [14].

We identified 74 articles describing hepatitis B and hepatitis E seroprevalence using blood donor specimens. Our inclusion criteria were limited to studies after 1990, excluding many hepatitis B serological studies with the objective to evaluate or modify existing blood donor screening guidelines (i.e., not residual specimens). Hepatitis E virus was discovered in 1980, and transfusion transmitted hepatitis E virus was first reported in 2004. Therefore, many articles on hepatitis E serology evaluated the need for blood donor screening (e.g., [33–35]). An increase in the number of hepatitis E serological surveys using residual specimens was observed from 2016-2019 (e.g., [36]). We attribute this increase to a rising interest in estimating population immunity due to the availability of the Hecolin hepatitis E vaccine that was licensed in China in 2011 and the need to understand the incidence of hepatitis E virus infection [37].

Metadata accompanying residual specimens, such as extended demographic meta-data (e.g., residence, socioeconomic status, occupation) and epidemiologic meta-data (e.g., vaccination status or infection history), are valuable to improve the interpretation of serological markers. For example, Murhekar et al. 2021 used stored samples from a dengue serosurvey linked to extended demographic data to estimate diphtheria seroprevalence by rural or urban residence and region of India [38]. Similarly, Yan et al. 2019 tested stored samples from a hepatitis B serosurvey to estimate hepatitis A seroprevalence. Because the original samples included demographic and epidemiologic meta-data about hepatitis A vaccine, they investigated the change in seroprevalence before and after vaccine introduction and compared vaccinated to unvaccinated individuals [39]. It would be useful to have a standard set of basic meta-data that are linked and reported on when residual specimens are used, acknowledging the ethical considerations and need for informed consent when collecting or abstracting additional data. Canada’s COVID-19 Immunity Task Force recommended a core set of meta-data to be collected, including demographics and history of vaccination or infection, that was used to standardize analyses across residual specimens collected during the early years of the COVID-19 pandemic [40].

Understanding how potential selection bias is handled allows readers to interpret the results and generalizability of the findings from residual specimens [41]. We were lenient in what we classified as exploring selection bias of residual specimens either in the design, analysis, or interpretation sections of the text because it was not always possible to extract the authors’ intentions. Most articles (77.5%) compared their results to other published estimates. If the estimates are comparable, this approach can lend some validity to the findings based on residual specimens but the study then provides less added value unless extended over a longer timeframe. This is particularly true if the comparison is to seroprevalence estimates from a representative sample in a neighboring area or similar time frame. We also considered the stratification of results as a method to control for differences in sample and target populations (e.g., by age, sex, or location) and ensure that each subgroup is represented in the analysis. Albeit the authors’ reason for stratification is generally unknown and it is likely some authors were simply interested in demographic-specific seroprevalence. However, design-phase methods of stratified subsampling, using inclusion and exclusion criteria as well as analytic methods of post-hoc weighting of results or analytic comparisons to alternative data sources are purposeful approaches to account for selection bias. These approaches are ideal but require appropriate meta-data to inform selection or weighting, or access to alternative data sources. For example, Choisy et al. 2019 used residual samples for clinical purposes stored at a Vietnamese national biobank to estimate measles seroprevalence. The authors directly addressed the potential for selection bias, subsampled by age, sex, and location, stratified the findings by these same demographic characteristics, and compared estimates to measles cases and measles vaccination coverage to interpret and validate results [42].

We identified three studies that directly compared seroprevalence estimates from residual clinical specimens to seroprevalence estimates from a population-based serosurvey to specifically address the utility of residual specimens; two estimated COVID-19 seroprevalence in the US [43,44] and one estimated seroprevalence to VPDs in Australia [45]. All three studies conducted intentional design-phase or analytic-phase approaches to address selection bias and found that seroprevalence estimates from residual specimens were comparable to those obtained from the population-based serosurvey. This gives credibility to the use of residual specimens in some settings and the ability to account for selection bias. We also identified 95 papers that used residual specimens to evaluate trends over time. Assuming the type and magnitude of selection bias stays constant across the period of interest, using residual specimens for this purpose can also overcome potential selection bias to obtain useful information.

Two-thirds of the articles used specimens from high-income countries, despite the larger burden of VPDs in low and middle-income countries. Scaling up of serosurveys using residual specimens may prove to be of greater value in resource limited settings, given that using residual specimens require less time, cost, and resource capacity compared to population-based serosurveys. For example, Kelly et al 2002 found that a serosurvey using residual clinical specimens was 11 times less costly than a population-based school serosurvey [54]. However, there remain important cost considerations after specimen collection regardless of the savings gained by using residual specimens. For example, health facilities may not have the capacity to store residual specimens collected for routine testing purposes. To use these specimens for additional testing, specimens need to be transported to another laboratory (e.g., research institute or district-level health facility) for storage until testing, along with linked data. This requires staff to process specimens and extract data as well as transportation to move specimens between facilities. Similarly, specimen repositories, such as those from past serosurveys, require sustained infrastructure to maintain frozen specimens.

In addition to the logistical considerations, there are ethical considerations to using specimens for a different purpose than originally intended. Much of the bioethics work related to residual specimens is focused on genomics or molecular disease surveillance, which have specialized considerations related to the types of data available and purposes (e.g., “readily identifiable” genetic information and tracking transmission networks) [46,47]. For serological surveys using residual specimens to develop population-level estimates of VPDs, information is typically presented as summary-level measures and ideally used by a public health agency to inform vaccination programs. Although this is generally accepted as benefiting the community, there are still important ethical considerations that are relevant for any research using residual specimens. These include understanding the population whose specimens were collected (e.g., whether they include vulnerable populations), the original purpose of the specimens, whether the individuals whose specimens were collected were aware of the potential for future research, if there were any formal consent or “opt-in/opt-out” requirements related to future research, and the policy and regulatory environment in the setting where the specimens were collected. Information about future research on residual specimens is typically provided during the consent process for research studies. We found many inconsistencies in how articles discussed permission to use residual specimens and ethical considerations. Many articles reported ‘broad consent’ was obtained at the time of specimen collection, although not all countries or settings allow this [48]. We identified only two articles that described reconsenting individuals and both involved contacting individuals or their caregivers to obtain consent for the additional testing [49,50]. How information about future research is communicated to blood or plasma donors or patients varies by setting. Studies using clinical specimens often described the specimens as being de-identified or anonymized; however, these terms may be used or interpreted differently, and policies and procedures related to these vary by country [51]. For example, in the United States, there are 18 specified identifiers that must be removed for the data to be considered “de-identified” [52]. Few of these identifiers are relevant for seroprevalence studies but data on age and geographic location are usually important, so special considerations to access these data may be needed depending on the research questions.

Our analysis identified articles that used residual specimens collected as part of serological surveillance systems. For example, the European Sero-Epidemiology Network (ESEN) is a network of European countries that aim to standardize serological surveillance for comparison across space and time. Some of the countries rely on residual specimens, while others use population-based serosurveys [53]. This supports the idea that serological surveillance systems can leverage residual specimens (e.g., diagnostic specimens). In turn, specimens from serosurveillance systems can be stored in biorepositories for future use. Serological surveillance systems can also be integrated, relying on multiplex technology to test for IgG antibodies to many antigens and pathogens simultaneously. The expansion of integrated serosurveillance to more low and middle-income settings is where there is highest value [16,54]. Integrated serological surveillance systems, particularly those that use multiplex assay technologies to simultaneously test for multiple pathogens, have the potential to improve the scale-up and utility of serological surveys, thus requiring less specimen volume [55]. Integrated serosurveillance can prospectively collect new specimens, as in the Netherlands [56], or take advantage of residual specimens, including from population based serosurveys for HIV [57] or malaria [58].

The design of a serosurvey using residual specimens varies by the VPD, target population, and research question of interest. Given the specified challenges in screening for articles that report the use of residual specimens and inconsistencies in how authors discussed the use of residual specimens, we recommend a standard set of descriptors for serological studies using residual specimens (Table 2). These recommendations are motivated by: 1) the need to detail the original specimen source to understand the potential for selection bias; 2) the ethics such that it is clear whether permissions were obtained and ethics of using residual specimens was considered; 3) documentation of broad learnings from serological surveys using residual specimens so that we continue to evaluate the limitations and potential strengths of these studies for the control of VPDs; and 4) the ability to use seroprevalence estimates for further analyses including comparisons across settings or meta-analyses.

**Table 2.** Recommendations on reporting studies using residual specimens.

| Component | Considerations |
| --- | --- |
| Study population | Details on the source of the specimens and description of the original population or study (including sample size and sociodemographic characteristics) to assess the generalizability of findings from residual specimens |
|  | Original reason or objective for specimen collection prior to storage |
|  | How residual specimens were sampled including inclusion and exclusion criteria (e.g., by time period, individual characteristics) |
| Ethics | Whether the individuals whose specimens were collected were aware of potential for future research; if there were any formal consent or “opt-in/opt-out” requirements related to future research; ethical approvals or waivers obtained, whether or not specimens were identifiable |
| Meta-data | How data were collected from the original population (including if part of a larger surveillance collection system) and what metadata were obtained, including:<br>• Age (e.g. DOB, age category)<br>• Sex/gender<br>• Geographic location (e.g. GPS, community, health facility catchment area, administrative 3 level)<br>• Vaccination history data<br>Any other available sociodemographic data. |
| Serological testing | Serological testing that was performed for the original purpose prior to being stored |
| Selection bias and generalizability | How the study handled and assessed selection biases in the residual specimen source population relative to the target population. This explanation is key to provide readers an understanding of how representative the findings may be of the target population. This could also include any potential limitations in the generalizability of the findings. |

There were several limitations in conducting this scoping review. Despite using a broad search strategy to capture the varied terms used to describe residual specimens, there may have been articles that published results from VPD serosurveys that used residual specimens missed by our search. For example, for many articles, particularly those using specimens from patients and blood donors, it was difficult to ascertain if the specimens were left over from clinical blood collection or diagnostic testing, or if they were purposefully collected from convenience populations for a serosurvey and therefore not residual specimens. Additionally, there was a class of articles that came from large national cross-sectional serosurveys in which it was difficult to determine the original purpose of the specimen collection to ascertain if the specimens were indeed residual (e.g., PIENTER or NHANES). There was a second class of articles from serosurveillance systems (e.g., ESEN) for which it was also challenging to determine if residual specimens were used. Ultimately, if there was no clear evidence the specimens were residual, then the article was excluded. This rule likely excluded articles that used residual specimens but did not explicitly describe them as such. Given only 10% of all articles met the screening criteria, there were challenges in narrowing the search strategy and screening. Lastly, among the 25 VPDs, 10 VPDs were studied by fewer than 1% of the included articles (Hib, Japanese encephalitis, tick-borne encephalitis, yellow fever, meningococcal meningitis, pneumococcus, rotavirus, tuberculosis, typhoid fever, and cholera). This low number likely reflects the fact that serological tests are not well-suited to study these pathogens [9,26], or that under-represented geographies in serological surveys are also where the burden of these pathogens are most prevalent.

## Conclusion

Serological surveys that rely on residual specimens have been used for decades. Their value to estimate seroprevalence to inform disease burden estimates (e.g., COVID-19 [59]), assess trends in infection, or risk factors for exposure (e.g., pandemic influenza H1N1 [60]) is especially noted during outbreaks of novel pathogens. During the COVID-19 pandemic when countries wanted rapid assessments of the prevalence of infection, vaccine effectiveness, waning immunity, and population susceptibility, residual specimens were a key source of serological data [61]. Serological surveys using residual specimens were also a valuable source to inform preventative strategies (e.g., measles [62]) or epidemiologically understand re-emergent outbreaks (e.g., measles [29]). The primary concern with residual specimens is whether they are representative of the target population. By reporting key parameters about study design and results, we can better evaluate the potential biases of studies using residual specimens to alleviate concerns of external validity or extract valuable information despite these biases. Ultimately, having a serosurveillance system in place that leverages residual specimens provides a platform to test for emerging pathogens, supporting pandemic preparedness initiatives [63,64].

## Data Availability

All data produced are available online at https://github.com/HopkinsIVAC/ScopingReview_SerosurveyResidual

## Acknowledgements

- Claire Twose, Welch Medical Library
- Natalya Kostandova, JHU for translating articles in Russian
- Berman Institute of Bioethics: Joseph Ali, Juli Bollinger, Debra Matthews

## Data and Supplemental Materials Available Online

https://github.com/HopkinsIVAC/ScopingReview_SerosurveyResidual

