## Supplemental Materials for "The use of residual blood specimens in seroprevalence studies for vaccine preventable diseases: A scoping review"

### Text S1: Protocol

**Review Question**

What are the characteristics, objectives, and considerations of serological studies that use residual blood specimens to measure population-level metrics?

**What are residual specimens within the scope of serological surveys?**

They are residual, remnant, surplus, existing, archived, or left-over serum that are serologically tested for reasons beyond the objective of the original sample collection.

Examples of residual sera include:

- Sera originally collected for other serosurveys, studies, or surveillance systems and stored.
- Sera originally collected for clinical purposes at a health facility or laboratory and stored.
- Sera originally collected for donation purposes and stored.

**Inclusion Criteria**

- Uses any residual serum samples for serological testing of infectious disease to measure immune status.
    - Note *any* means at least some of the serum samples are residual.
    - Residual serum samples for serological testing are remnant, surplus, existing, archived, or left-over serum that are serologically tested for reasons beyond the objective of the original sample collection.
    - The following are a list of diagnostic tests that measure immune status (i.e., not acute infection):
        - neutralization tests (NTs) - plaque reduction neutralization tests
        - hemagglutinin-inhibition (HI)
        - IgG immunoassays - enzyme immunoassay (EIA), enzyme-linked immunosorbent assay (ELISA), florescent immunoassays (FIA), radio-immunoassays (RIA), chemiluminescence immunoassays
        - Multiplex
        - Rapid test IgG
- Serological testing related to the 25 pathogens of interest (i.e., vaccine preventable diseases except for rabies, anthrax, and smallpox). The following are the pathogens of interest:
    - *Vibrio cholerae* (cholera), dengue, *Corynebacterium diptheriae* (diphtheria), hepatitis A (HAV), hepatitis B (HBV), hepatitis E (HEV), *Haemophilus influenza* type b (Hib), human papillomavirus (HPV), influenza, Japanese encephalitis, measles, *Neisseria meningitidis* (meningococcal meningitis), mumps, *Bordetella pertussis* (pertussis), *Streptococcus pneumoniae* (pneumococcal), poliovirus (poliomyelitis), rotavirus, rubella, SARS-CoV-2 (COVID-19), *Clostridium tetani* (tetanus), tick-borne encephalitis, *Mycobacterium tuberculosis* (tuberculosis), *Salmonella* Typhi (typhoid fever), varicella-zoster (herpes zoster/shingles & varicella/chickenpox), yellow fever
- Peer-reviewed literature, grey literature, and pre-prints
- English, French, Spanish, Portuguese, Italian, Russian, Chinese language articles published between January 1, 1990 and August 22, 2022

**Exclusion Criteria**

- Serological testing of pathogens of non-interest: all non-VPDs (e.g., HIV, Hepatitis C, malaria, etc), and three VPDs (rabies, anthrax, smallpox)

- Use residual sera samples for serological testing of infectious disease to test for primary or acute infection only. The following are a list of diagnostic tests that measure acute infection (i.e., not immune status):
    - nucleic acid detection assays (e.g., PCR)
    - IgM immunoassays
    - Complement fixation test (CFT)
    - Point-of-care (POC) tests
- Use residual sera samples to evaluate diagnostic tests or inform diagnostic tests laboratory protocols
- Use residual sera samples as a control (or comparison) group to a non-residual population of interest
- Use residual samples to modify existing blood donor screening guidelines/protocols; *existing* meaning we exclude studies that use residual specimens to determine *how to screen* for an already routinely screened pathogen (note: this differs from studies that are attempting to determine *whether to screen* for a new pathogen, which would be included).
- Biological samples are not blood sera. The following are a list of biological specimens that are not a specimen of interest:
    - placental/umbilical cord sera, cerebrospinal fluid sample, oral fluid sample, ocular specimens/fluid, tissue sample
- Animal studies and studies in other non-human specimens
- Editorials, letters, commentaries, narrative reviews, conference abstracts

**Databases**

- PubMed
- Scopus
- Embase
- Cochrane
- WHO IRIS

**Search Terms**

<u>PubMed (first search conducted 6/23/2021; second search conducted on 8/22/2022)</u>
("measles"[Mesh] OR "measles virus"[Mesh] OR "measles"[tiab] OR "sars-cov-2"[MeSH Terms] OR "sars-cov-2"[All Fields] OR "sars cov 2"[All Fields] OR "sars-cov-2"[tiab] OR "whooping cough"[Mesh] OR "pertussis"[tiab] OR "rubella"[Mesh] OR "rubella"[tiab] OR "papillomaviridae"[Mesh] OR "human papillomavirus"[tiab] OR "influenza, human"[Mesh] OR "cholera"[Mesh] OR "cholera"[tiab] OR "neisseria meningitidis"[Mesh] OR "meningococcal infections"[Mesh] OR "diphtheria"[Mesh] OR "diptheria"[tiab] OR "mumps"[Mesh] OR "mumps"[tiab] OR "tetanus"[Mesh] OR "tetanus toxoid"[Mesh] OR "tetanus"[tiab] OR "Hepatitis A"[Mesh] OR "Hepatitis B"[Mesh] OR "Hepatitis E"[Mesh] OR "hepatitis"[tiab] OR "Tuberculosis"[Mesh] OR "Pneumococcal Infections"[Mesh] OR "Typhoid Fever"[Mesh] OR "Poliomyelitis"[Mesh] OR "haemophilus influenzae type b"[mesh] OR "Herpes Zoster"[Mesh] OR "Herpesvirus 3, Human"[Mesh] OR "varicella"[tiab] OR "Rotavirus"[Mesh] OR "Rotavirus Infections"[Mesh] OR "rotavirus"[tiab] OR "Yellow Fever"[Mesh] OR "Encephalitis, Japanese"[Mesh] OR "Dengue"[Mesh] OR "dengue"[tiab] OR "Encephalitis, Tick-Borne"[Mesh])
AND
("serology"[MeSH Terms] OR "serology"[All Fields] OR "serologic tests"[MeSH Terms] OR ("serologic"[All Fields] AND "tests"[All Fields]) OR "serologic tests"[All Fields] OR "serologies"[All Fields] OR "seroepidemiology"[All Fields] OR "seroepidemiologic"[All Fields] OR “seroepidemiologic studies”[Mesh] OR "seroepidemiological"[All Fields] OR "seroprevalence"[All Fields] OR "seropositive"[All Fields] OR "seroconversion"[All Fields] OR "sero-conversion"[All Fields] OR

("serologic"[All Fields] AND "surveillance"[All Fields]) OR ("serological"[All Fields] AND "surveillance"[All Fields]) OR "serosurvey"[All Fields])
AND
((“residual"[All Fields] OR "stored”[All Fields] OR "leftover"[All Fields] OR "existing"[All Fields] OR "remnant"[All Fields] OR “archived”[All Fields] OR "convenience"[All Fields]) AND ("serum"[All Fields] OR "sera"[All Fields] OR "sample*"[All Fields] OR "specimen*"[All Fields])
OR
“biorepository”[All Fields] OR “biobank*”[All Fields] OR ("serum"[All Fields] AND "bank*"[All Fields]) OR "blood donor*"[All Fields] OR “population-based”[all fields] OR “general population”[all fields])
AND 1990/01/01:2021/06/23[dp]

Embase (first search conducted 6/23/2021; second search conducted on 8/22/2022)
('measles'/exp OR 'measles virus'/exp OR measles:ab,ti OR measles OR 'sars-cov-2'/exp OR 'sars-cov-2':ab,ti OR 'whooping cough'/exp OR pertussis:ab,ti OR 'rubella'/exp OR rubella:ab,ti OR 'papillomaviridae'/exp OR 'human papillomavirus':ab,ti OR 'influenza next/2 human' OR influenza OR 'cholera'/exp OR cholera:ab,ti OR 'neisseria meningitidis'/exp OR 'meningococcal infections'/exp OR 'diphtheria'/exp OR diphtheria:ab,ti OR 'mumps'/exp OR mumps:ab,ti OR 'tetanus'/exp OR 'tetanus toxoid'/exp OR tetanus:ab,ti OR 'hepatitis a'/exp OR 'hepatitis b'/exp OR 'hepatitis e'/exp OR hepatitis:ab,ti OR 'tuberculosis'/exp OR 'pneumococcal infections'/exp OR 'typhoid fever'/exp OR 'poliomyelitis'/exp OR 'haemophilus influenzae type b'/exp OR 'herpes zoster'/exp OR 'herpesvirus 3 next/2 human' OR varicella:ab,ti OR 'rotavirus'/exp OR 'rotavirus infections'/exp OR rotavirus:ab,ti OR 'yellow fever'/exp OR 'japanese encephalitis'/exp OR 'dengue'/exp OR dengue:ab,ti OR 'tick-borne encephalitis'/exp)
AND
('serology'/exp OR serology OR 'serologic tests'/exp OR (serologic NEAR/3 tests) OR 'serolog*' OR 'seroepidemiolog*' OR (‘seroepidemiolog*’ NEAR/3 studies) OR 'seroprevalence'/exp OR seroprevalence OR 'seropositiv*' OR seropositive OR 'seroconversion'/exp OR 'sero conversion' OR (serologic* NEAR/3 surveillance) OR 'serosurvey' OR serosurvey)
AND
(((residual OR stored OR leftover OR existing OR remnant OR archived OR convenience) NEAR/3 (serum OR sera OR sample* OR specimen*)) OR 'biorepository'/exp OR biorepository OR 'biobank*' OR (serum NEXT/2 bank*) OR 'blood donor*' OR ‘population-based’ OR ‘general population’)
AND [1990-2021]/py

Scopus (first search conducted 6/24/2021; second search conducted on 8/22/2022)
TITLE-ABS-KEY("measles" OR "measles virus" OR "sars-cov-2" OR "sars cov 2" OR "whooping cough" OR "pertussis" OR "rubella" OR "papillomaviridae" OR "human papillomavirus" OR "influenza, human" OR "cholera" OR "neisseria meningitidis" OR "meningococcal infections" OR "diphtheria" OR "mumps" OR "tetanus" OR "tetanus toxoid" OR "Hepatitis A" OR "Hepatitis B" OR "Hepatitis E" OR "hepatitis" OR "Tuberculosis" OR "Pneumococcal Infection*" OR "Typhoid Fever" OR "Poliomyelitis" OR "haemophilus influenzae type b" OR "Herpes Zoster" OR "Herpesvirus 3, Human" OR "varicella" OR "rotavirus" OR "Yellow Fever" OR "Japanese Encephalitis" OR "dengue" OR "Tick-Borne Encephalitis")
AND
ALL("serology" OR "serolog* tests" OR ("serologic" W/3 "tests") OR "serolog*" OR "seroepidemiolog*" OR “seroepidemiolog* studies” OR "seroprevalence" OR "seropositive" OR "seroconversion" OR "sero-conversion" OR ("serologic*" W/3 "surveillance") OR "serosurvey")
AND

ALL((“residual" OR "stored” OR "leftover" OR "existing" OR "remnant" OR “archived” OR "convenience") AND ("serum" OR "sera" OR "sample*" OR "specimen*") OR “biorepository” OR “biobank*” OR "serum bank*" OR "blood donor*" OR “population-based” OR “general population”) AND PUBYEAR > 1990

Cochrane (first search conducted 6/28/2021; second search conducted on 8/22/2022)
("measles" OR "measles virus" OR "sars-cov-2" OR "sars cov 2" OR "whooping cough" OR "pertussis" OR "rubella" OR "papillomaviridae" OR "human papillomavirus" OR "influenza, human" OR "cholera" OR "neisseria meningitidis" OR "meningococcal infections" OR "diphtheria" OR "mumps" OR "tetanus" OR "tetanus toxoid" OR "Hepatitis A" OR "Hepatitis B" OR "Hepatitis E" OR "hepatitis" OR "Tuberculosis" OR "Pneumococcal Infection*" OR "Typhoid Fever" OR "Poliomyelitis" OR "haemophilus influenzae type b" OR "Herpes Zoster" OR "Herpesvirus 3, Human" OR "varicella" OR "rotavirus" OR "Yellow Fever" OR "Japanese Encephalitis" OR "dengue" OR "Tick-Borne Encephalitis") in Title Abstract Keyword
AND ("serology" OR "serolog* tests" OR "serologic tests" OR "serolog*" OR "seroepidemiolog*" OR “seroepidemiolog* studies” OR "seroprevalence" OR "seropositive" OR "seroconversion" OR "sero-conversion" OR "serologic* surveillance" OR "serosurvey") in All Text
AND ((“residual" OR "stored” OR "leftover" OR "existing" OR "remnant" OR “archived” OR "convenience") AND ("serum" OR "sera" OR "sample*" OR "specimen*") OR “biorepository” OR “biobank*” OR "serum bank*" OR "blood donor*" OR “population-based” OR “general population”) in All Text

**Search strategy**
Screening was first conducted through an online PubMed search for English-language and French-language literature published between January 1990 and June 2021. Between June 2021 and July 2021, we searched several electronic databases with reference to the expanded Medical Subject Headings (MeSH) thesaurus, using search terms for vaccine-preventable diseases (VPDs), serologic testing, and residual samples. The databases included the following: PubMed, Scopus, Embase, Cochrane, and the WHO iris database. Additional studies were identified through manual searches of the reference lists of identified papers including systematic reviews and papers reviewing biobank utility. Suggestions from experts in the field and investigations from two international organizations (CDC and WHO) were consulted.

**Screening Strategy**
After deleting duplicates, two members of the literature review group systematically screened the title and abstract of papers identified in the electronic searches for the inclusion and exclusion criteria using Covidence. Those that met the criteria or have remaining uncertainty for the criteria were read in full text. Full text screening was done by two reviewers with all disagreements resolved through discussion and arbitration with a third reviewer. Prior to the final analysis, the search was re-run (August 2022) to include studies published while reviewers conducted the initial screening.

**Condition or domain being studied**
The scoping review aims to describe the uses of residual sera samples in serological testing studies of VPDs.

**Participants/Population**
Humans of all ages, regardless of location and characteristics.

**Main Outcome(s)**
The purpose for using residual samples in the study, i.e., seroprevalence of a pathogen. The following will be considered to inform the development of key themes describing the use of residual sera samples:

- VPD of interest
- Country and region from where samples were collected
- Collection and storage site of specimen
- Specimen type (venous or finger-prick)
- Sample size
- Original use for the samples
- Study year(s)
- Year(s) of original sample collection
- Population from where samples were collected
- Serological tests that were performed
- Statement on permissions and ethical considerations associated with using residual sera
- Meta-data of the residual sample
- Objectives of the paper
- If and how authors talk about bias of residual specimens
- If collection was part of a larger serological surveillance system

**Data Extraction**

A standardized data extraction form will be developed in Kobo Toolbox and piloted on 5-10 known publications. Any issues will be addressed, and the final version will be used. One reviewer will conduct data extraction. A second reviewer will be consulted if questions arise about adherence to inclusion and exclusion criteria or the data extraction process. The following data will be extracted: publication details, type of study, study design and methods, results, and methodological quality items according to the type of study. The main outcomes will be extracted from publications in the following manner:

Identification

- Publication year
- Country and region: codelist [country codes]
- Study year(s): codelist [1980 – 2022]
- Year(s) of original sample collection: dropdown [1900 – 2022]

Population

- Population that sample was collected from: codelist [general population, pregnant women, blood donors, plasma donors, patient population, school children, injection drug users, non-injecting drug users, military, MSM, homeless population, healthcare workers, tattoo artists, indigenous people, migrants, inmates/incarcerated individuals, university students, not specified]
- Details of the original population: free text
- Original use for the samples: codelist [blood donation, plasma donation, seroprevalence study, diagnostic sample, other/non-serological survey]
- Collection site of original sample: codelist [household, school, hospital/clinic, blood or plasma donation site, diagnostic center/laboratory, homeless shelter, prison, unknown]
- Storage site of residual specimens: codelist [research biorepository, blood/plasma donation center biorepository, diagnostic center/laboratory biorepository, unspecified biorepository]
- Total sample size of the whole analysis: free text
- Sample size of residual specimens: free text
- Specimen type: codelist [fingerprick, venous]

Methods

- Paper objective: codelist [to describe population seropositivity, to describe seroprevalence among a specific clinical subpopulation, to evaluate the need for blood donor screening, to describe antibody kinetics following vaccination (e.g., vaccine effectiveness), to describe antibody kinetics following natural infection (e.g., immunogenicity of infection), to estimate infection rates or understand transmission dynamics, to compare the use of residual samples to population-based samples, to identify risk factors of seropositivity, to evaluate outcomes of seropositivity, to evaluate changes in seropositivity overtime, to evaluate cross-reactivity, to describe viral dynamics, to compare population seropositivity between countries or international regions]

- VPD of interest is main objective or subobjective: binary [yes, no]
- Residual sample came from a national sero-surveillance system or a national serial cross-sectional serosurvey: binary [yes, no]
    - If yes: name the national sero-surveillance system or a national serial cross-sectional serosurvey
- Inclusion criteria for residual samples: free text
- Exclusion criteria for residual samples: free text
- Serological tests that were performed: codelist [enzyme-linked immunosorbent assay or enzyme-based immunoassay (ELISA or EIA), hemagglutination inhibition (HI) assay, neutralization test (NT or PRNT), chemiluminescence immunoassay (CLIA), time-resolved fluoroimmunoassay (TR-FIA), radioimmunoassay/immunofluorescence technique (IF), multiplex immunoassay, lateral flow rapid test, epitope-blocking assay, isoelectric focusing, Western blot, latex agglutination test, protein microarray, fluorescent-antibody-to-membrane-antigen (FAMA) test, toxin binding inhibition assay]
- VPD of interest: codelist [cholera, dengue, diptheria, HAV, HBV, HEV, Hib, HPV, influenza, Japanese encephalitis, measles, mumps, Neisseria meningitis, pertussis, pneumococcal, polio, rotavirus, rubella, Sars-CoV-2, tetanus, tick-borne encephalitis, tuberculosis, typhoid, varicella/herpes zoster, yellow fever]
- Meta-data linked to residual samples and used in the analysis: codelist [basic demographic data (age, sex, race, ethnicity), extended demographic data (e.g., residence, occupation, SES), epidemiologic data on VPD of interest (e.g., vaccination status, history of infections), epidemiologic data not on VPD of interest (e.g., history or current infection status of other diseases, biomarkers), none]

Quality Assessment

- Authors included a statement about ethical considerations associated with retesting specimens: codelist [approved by an ethics committee/board, broad consent given at time of specimen collection, consent given in retrospect, received waiver on re-consenting, samples deidentified/anonymized, exempt as public health surveillance, did not include a statement]
- Acknowledgement of bias from residual samples: codelist [no mention of bias, acknowledged bias, residual sample not biased (because the target population of the residual sample is the same as the target population of the original sample)]
- Handling of bias: codelist [conducted stratified subsampling, used inclusion/exclusion criteria for the samples, conducted stratified analysis, weighted final results, compared to alternative data source for "validity", compared to other published estimates, tested samples multiple times for the same pathogen, conducted sensitivity analysis, did not account for bias/none of the above]

**Strategy for data synthesis**

Data synthesis will be descriptive. The scoping review will be a narrative on the uses of residual samples in serologic studies. VPDs will be categorized based on primary mode of transmission for data synthesis (airborne, vector-borne, food- or water-borne, human physical contact, other physical contact). The data synthesis will include enumeration of the main outcomes and presented in tables, maps, and graphs.

**Text S2: Preferred Reporting Items for Systematic reviews and Meta-Analyses extension for Scoping Reviews (PRISMA-ScR) Checklist**

| SECTION | ITEM | PRISMA-ScR CHECKLIST ITEM | REPORTED ON PAGE # |
|---|---|---|---|
| **TITLE** | | | |
| Title | 1 | Identify the report as a scoping review. | See Title Page |
| **ABSTRACT** | | | |
| Structured summary | 2 | Provide a structured summary that includes (as applicable): background, objectives, eligibility criteria, sources of evidence, charting methods, results, and conclusions that relate to the review questions and objectives. | See Abstract |
| **INTRODUCTION** | | | |
| Rationale | 3 | Describe the rationale for the review in the context of what is already known. Explain why the review questions/objectives lend themselves to a scoping review approach. | Reported in introduction section |
| Objectives | 4 | Provide an explicit statement of the questions and objectives being addressed with reference to their key elements (e.g., population or participants, concepts, and context) or other relevant key elements used to conceptualize the review questions and/or objectives. | Reported in introduction section |
| **METHODS** | | | |
| Protocol and registration | 5 | Indicate whether a review protocol exists; state if and where it can be accessed (e.g., a Web address); and if available, provide registration information, including the registration number. | Reported in method section that protocol exists in supplementary materials (Text S1) |
| Eligibility criteria | 6 | Specify characteristics of the sources of evidence used as eligibility criteria (e.g., years considered, language, and publication status), and provide a rationale. | Summarized in methods section and detailed in protocol (Text S1) |
| Information sources* | 7 | Describe all information sources in the search (e.g., databases with dates of coverage and contact with authors to identify additional sources), as well as the date the most recent search was executed. | Summarized in methods section and detailed in protocol (Text S1) |
| Search | 8 | Present the full electronic search strategy for at least 1 database, including any limits used, such that it could be repeated. | Included in the protocol (Text S1) |
| Selection of sources of evidence† | 9 | State the process for selecting sources of evidence (i.e., screening and eligibility) included in the scoping review. | Summarized in methods section and detailed in protocol (Text S1) |
| Data charting process‡ | 10 | Describe the methods of charting data from the included sources of evidence (e.g., calibrated forms or forms that have been tested by the team before their use, and whether data charting was done independently or in duplicate) and any processes for obtaining and confirming data from investigators. | Summarized in methods section and detailed in data extraction survey (Text S3) |

| | | | |
|---|---|---|---|
| Data items | 11 | List and define all variables for which data were sought and any assumptions and simplifications made. | Summarized in methods section and detailed in data extraction survey (Text S3 and S4) and supplementary material (Table S1-S3) |
| Critical appraisal of individual sources of evidence§ | 12 | If done, provide a rationale for conducting a critical appraisal of included sources of evidence; describe the methods used and how this information was used in any data synthesis (if appropriate). | Articles were not critically appraised; stated in the methods section |
| Synthesis of results | 13 | Describe the methods of handling and summarizing the data that were charted. | Reported in the methods section and supplementary material (Table S1-S3) |
| **RESULTS** | | | |
| Selection of sources of evidence | 14 | Give numbers of sources of evidence screened, assessed for eligibility, and included in the review, with reasons for exclusions at each stage, ideally using a flow diagram. | Summarized in results section and detailed in Figure 1 |
| Characteristics of sources of evidence | 15 | For each source of evidence, present characteristics for which data were charted and provide the citations. | Summary of the articles organized by VPD is included in supplementary materials (Table S4) |
| Critical appraisal within sources of evidence | 16 | If done, present data on critical appraisal of included sources of evidence (see item 12). | Not applicable |
| Results of individual sources of evidence | 17 | For each included source of evidence, present the relevant data that were charted that relate to the review questions and objectives. | Summary of the articles organized by VPD is included in supplementary materials (Table S4) |
| Synthesis of results | 18 | Summarize and/or present the charting results as they relate to the review questions and objectives. | Reported throughout the results section, Table 1, Figure 3-6, and supplementary materials (Text S5, Figures S1-S3, Tables S5-S9) |
| **DISCUSSION** | | | |
| Summary of evidence | 19 | Summarize the main results (including an overview of concepts, themes, and types of evidence available), link to the review questions and objectives, and consider the relevance to key groups. | Reported throughout the discussion section |
| Limitations | 20 | Discuss the limitations of the scoping review process. | Reported in discussion section |
| Conclusions | 21 | Provide a general interpretation of the results with respect to the review questions and objectives, as well as potential implications and/or next steps. | Reported in discussion section |
| **FUNDING** | | | |
| Funding | 22 | Describe sources of funding for the included sources of evidence, as well as sources of | Reported in funding section |

| | | |
|---|---|---|
| | | funding for the scoping review. Describe the role of the funders of the scoping review. |

JBI = Joanna Briggs Institute; PRISMA-ScR = Preferred Reporting Items for Systematic reviews and Meta-Analyses extension for Scoping Reviews.
* Where *sources of evidence* (see second footnote) are compiled from, such as bibliographic databases, social media platforms, and Web sites.
† A more inclusive/heterogeneous term used to account for the different types of evidence or data sources (e.g., quantitative and/or qualitative research, expert opinion, and policy documents) that may be eligible in a scoping review as opposed to only studies. This is not to be confused with *information sources* (see first footnote).
‡ The frameworks by Arksey and O'Malley (6) and Levac and colleagues (7) and the JBI guidance (4, 5) refer to the process of data extraction in a scoping review as data charting.
§ The process of systematically examining research evidence to assess its validity, results, and relevance before using it to inform a decision. This term is used for items 12 and 19 instead of "risk of bias" (which is more applicable to systematic reviews of interventions) to include and acknowledge the various sources of evidence that may be used in a scoping review (e.g., quantitative and/or qualitative research, expert opinion, and policy document).

*From:* Tricco AC, Lillie E, Zarin W, O'Brien KK, Colquhoun H, Levac D, et al. PRISMA Extension for Scoping Reviews (PRISMAScR): Checklist and Explanation. Ann Intern Med. 2018;169:467–473. doi: 10.7326/M18-0850.

### Text S3: Kobo Toolbox Data Extraction Survey

**Residual Sera Scoping Review – Data Extraction Questions**

1. Title of study
2. Last name of first author
3. Publication year
4. Covidence number
5. Year(s) that study was conducted: [1900 – 2022]
6. Study country(s): [list country codes]
7. Is a VPD the main objective of the research paper? Yes / No.
8. Did the residual sample come from a national sero-surveillance system or a national serial cross-sectional serosurvey? Yes / No. If Yes, specify.
9. Population that the residual sample was collected from:
   - General population
   - Pregnant women
   - Blood donors
   - Plasma donors
   - Patient population
   - School children
   - Injection drug users
   - Non-injecting drug users
   - Military
   - MSM
   - Homeless population
   - Healthcare workers
   - Tattoo artists
   - Indigenous people
   - Migrants
   - Inmates/Incarcerated individuals
   - University students
   - Not specified
   - Other: specify
10. Provide details of the original population.
11. Original use for samples:
    - Blood donation
    - Plasma donation
    - Serosurvey
    - Diagnostic sample (including routine testing and chronic patients)
    - Other/non-serological survey
    - Other: specify
12. Did the residual population have inclusion criteria? Yes / No. If Yes, provide details.
13. Did the residual population have exclusion criteria? Yes / No. If Yes, provide details.
14. Collection site of residual specimens:
    - Household

School
Hospital/clinic
Blood donation site
Plasma donation site
Diagnostic center/laboratory
Homeless shelter
Prison
Unknown
Other: specify

15. Storage site of residual specimens:
    Unspecified biorepository
    Research biorepository
    Blood/plasma donation center biorepository
    Diagnostic center/laboratory biorepository

16. Individual meta-data linked to residual samples and used in the analysis:
    Basic demographic data (age, sex, race, ethnicity)
    Extended demographic data (e.g., residence, occupation, SES)
    Epi data on VPD of interest (e.g., vaccination status, history of infections)
    Epi data not on VPD of interest (e.g., history or current infection status of other diseases, biomarkers)
    None

17. Did the authors include a statement about ethical considerations with retesting specimens?
    Yes, approved by an ethics committee/board
    Yes, broad consent given at time of specimen collection
    Yes, consent given in retrospect
    Yes, received waiver on re-consenting
    Yes, samples de-identified/anonymized
    Yes, exempt as public health surveillance
    Yes, other: specify
    No

18. Total sample size of the study

19. Sample size of residual specimens

20. Sample size of the original sample from which residual sample was pulled

21. Specimen type:
    Fingerprick
    Venous

22. Serological tests performed:
    Neutralization test
    Hemagglutination inhibition assay
    Enzyme-linked immunosorbent assay or enzyme-based immunoassay (ELISA or EIA)
    Chemiluminescence immunoassay (CLIA)
    Time-resolved fluoroimmunoassay (TR-FIA)
    Radioimmunoassay, immunofluorescence technique (IF)
    Multiplex immunoassay
    Lateral flow immunoassay (rapid test)
    Epitope-blocking assay using monoclonal antibodies
    Isoelectric focusing

Immunoblot (Western blot)
Latex agglutination (LA) test
Protein microarray (PA)
Fluorescent-Antibody-to-Membrane-Antigen (FAMA) test
Toxin binding inhibition assay
Other: specify

23. VPD(s) of interest:
    Cholera
    Dengue
    Diphtheria
    Hepatitis A (HAV)
    Hepatitis B (HBV)
    Hepatitis E (HEV)
    Hemophilus influenza (Hib)
    Human Papillomavirus (HPV)
    Influenza
    Japanese Encephalitis
    Measles
    Mumps
    Neisseria Meningitis (meningococcal meningitis)
    Pertussis (whooping cough)
    Pneumococcal
    Poliomyelitis (polio)
    Rotavirus
    Rubella
    Sars-CoV-2 (COVID-19)
    Tetanus
    Tick-borne Encephalitis
    Tuberculosis
    Typhoid (Salmonella Typhi or typhoid fever)
    Varicella/Herpes Zoster (shingles & chicken pox)
    Yellow Fever

24. Acknowledgement of bias from residual samples:
    No mention of bias
    Acknowledged bias
    Residual sample not biased (b/c the target population of the residual sample is the same as the target population of the original sample)

25. Exploring bias of residual samples:
    Conducted stratified subsampling
    Used inclusion/exclusion criteria for the samples to address the bias
    Conducted stratified analysis (results)
    Weighted final results
    Compared it to alternative data source for "validity"
    Compared to other published estimates (discussion)
    Did not account for bias/none of the above
    Conducted sensitivity analysis

26. Paper objective (identified in title/abstract):
    to describe population seropositivity
    to describe seroprevalence among a specific clinical subpopulation

to evaluate the need for blood donor screening (e.g., HEV)
to describe antibody kinetics following vaccination (e.g., vaccine effectiveness)
to describe antibody kinetics following natural infection (e.g., immunogenicity of infection)
to estimate infection rates or understand transmission dynamics
to compare the use of residual samples to population-based samples
to identify risk factors of seropositivity
to evaluate outcomes of seropositivity
to evaluate changes in seropositivity overtime
to evaluate cross-reactivity
to describe viral dynamics
to compare population seropositivity between countries or international regions
Other: specify

27. Other comments

### Text S4: Defining Serological Surveillance System

In question #8 above of the data extraction tool, we asked "Did the residual sample come from a national sero-surveillance system or a national serial cross-sectional serosurvey? Yes / No. If Yes, specify." We began with a short list of surveillance sero-surveillance systems and national serial cross-sectional serosurveys we were aware of. Then we added onto the list as data extraction continued. In the end we delineated the following:

1. Unspecified serial cross-sectional survey
2. Unspecified sero-surveillance system
3. NHANES (USA)
4. PIENTER (Netherlands)
5. ESEN (European) Studies
6. KiGGS (Germany)
7. NCIRS (Australia)
8. HPA sero-surveillance/seroepidemiology unit (England) - formerly PHLS Serological Surveillance Programme
9. UK biobank cohort
10. Vietnam sero-surveillance (serum biobank project)
11. CIRN (Canada)
12. ID Surveillance Center or National Serum Reference Bank (SRB) from National Institute of Infectious Diseases (Japan)
13. Israel national serum bank by Center for Disease Control (ICDC)
14. Blood and Organ Transmissible Infectious Agents (BOTIA) project
15. U.S. Defense Medical Surveillance System (DMSS) and Department of Defense Serum Repository
16. Hungarian serial cross-sectional sero-surveillance (National Centre of Epidemiology)
17. Finnish Maternity Cohort (FMC)
18. Singapore National Health Survey (NHS)
19. Iceland Maternity Cohort
20. National Health and Nutrition Survey (ENSANUT) Mexico
21. German Health Interview and Examination Survey for Adults (DEGS)
22. KNHANES (Korea)
23. National Pediatric Seroprevalence Survey (NPSS) Singapore
24. JANUS (Norway) cancer registry
25. GUMCAD STI Surveillance System (England)
26. German National Nutrition Survey (NVS)
27. Retrovirus Epidemiology Donor Study (REDS) Allogenic Donor Recipient (RADAR) repository
28. Swiss HIV Cohort Study (SHCS)
29. Israel Defense Force Health Surveillance Bank
30. US National Cancer Institute (NCI) cohort consortium
31. Diabetomobile (Germany)
32. Other

### Text S5: Supplemental Results

*Serosurvey objectives*

Within the article abstract or summary, researchers identified one or more of twelve study objectives delineated in Table S2. Figure 3 shows the percent of studies that report each objective overall and by the top 6 VPDs. Most articles (80.5%) specified at least one objective was to describe population seroprevalence (e.g., Ekong et al 2022 estimated dengue seroprevalence in Nigeria using stored specimens originally tested for acute brucellosis or malaria infection [7]). Over half of articles that did not have this objective described seroprevalence among a specific clinical subpopulation (e.g., Toyoda et al. 2008 used stored serum samples from hemophilia patients to assess the prevalence of HEV antibody in Japanese patients with hemophilia [213]) (Table S5). Other objectives were to identify risk factors of seropositivity (33.3%) (e.g., Chin et al 1991 evaluated age, sex, and socio-economic risk factors of hepatitis A seropositivity in Hong Kong by using residual specimens originally tested for hepatitis B [55]), estimate parameters relating to magnitude or timing of infection such as incidence, prevalence, force of infection (17.6%) (e.g., Routledge et al 2022 tested for COVID-19 seroprevalence using residual specimens from blood draws to estimate the probability of prior infection among geographic regions in San Francisco, USA [534]), and/or evaluate changes in seroprevalence over time (15.8%). A significantly larger proportion of influenza, HPV, and Hepatitis E studies evaluate cross-reactivity, evaluate clinical outcomes of seropositivity, and evaluate the need for blood donor screening, respectively (Figure 3).

*Sources of residual specimens*

The main source of residual specimens for all articles as well as those studying COVID-19, hepatitis B, HPV, influenza, and measles was from patient populations (Figure S1). However, hepatitis E articles, used blood and plasma donor samples more than patient samples (Figure S1). Residual blood from general populations were the second or third most used source of specimens for all VPD groups, particularly for HPV and influenza (Figure S1). As defined in Table S1, we categorized specimens from general populations if they were representative of a country, region or other geographic unit (e.g., Ang et al. 2015 tested residual specimens for dengue seropositivity that were originally collected as part of Singapore's population-based National Health Survey to determine the prevalence of chronic conditions among adults [24]) or not otherwise specified by other source populations such as vaccine trials or cohort studies (e.g., Thiry et al. 2002 used specimens from a serum bank collected from Belgium volunteers in a vaccine trial to test for varicella zoster virus seropositivity [25]). The proportion of articles using samples from vulnerable populations and drug users was higher for hepatitis B.

Over time, specimens collected from patients for routine or specialized testing made up the highest proportion of original populations per year between 2004 to 2018 (Figure S2A). Before 2004, blood and plasma donors were equal or greater in proportion than patients, but after 2018, blood and plasma donors were used in comparable amounts to patients (Figure S2A). Over time, clinical testing and blood or plasma donations were the leading purposes for original specimen collection (Figure S2B). Serosurveys as a purpose for original data collection are consistent over time (Figure S2B).

*Time since collection*

Articles tended to use recently collected specimens, as the mean difference in years between publication and original sample collection was 6.5 years (Figure S3A). Eighty percent and ninety-four percent of articles used specimens collected within 10 or 20 years of publication, respectively. The mean number of years between data collection and publication for COVID-19 was 1.7 years (Figure S3B).

**Figure S1. Bar plot displaying percent of studies by original sample population for all VPDs and for the six most studied VPDs (N = 601 studies).** Note, the categories below are not mutually exclusive meaning that one study could have conducted testing for multiple pathogens and have had multiple original sample population. Also note that occupational populations include military, healthcare workers, farmers, and other occupational cohorts, and vulnerable populations include indigenous people, inmates/incarcerated individuals, homeless population, men who have sex with men (MSM), and migrants.

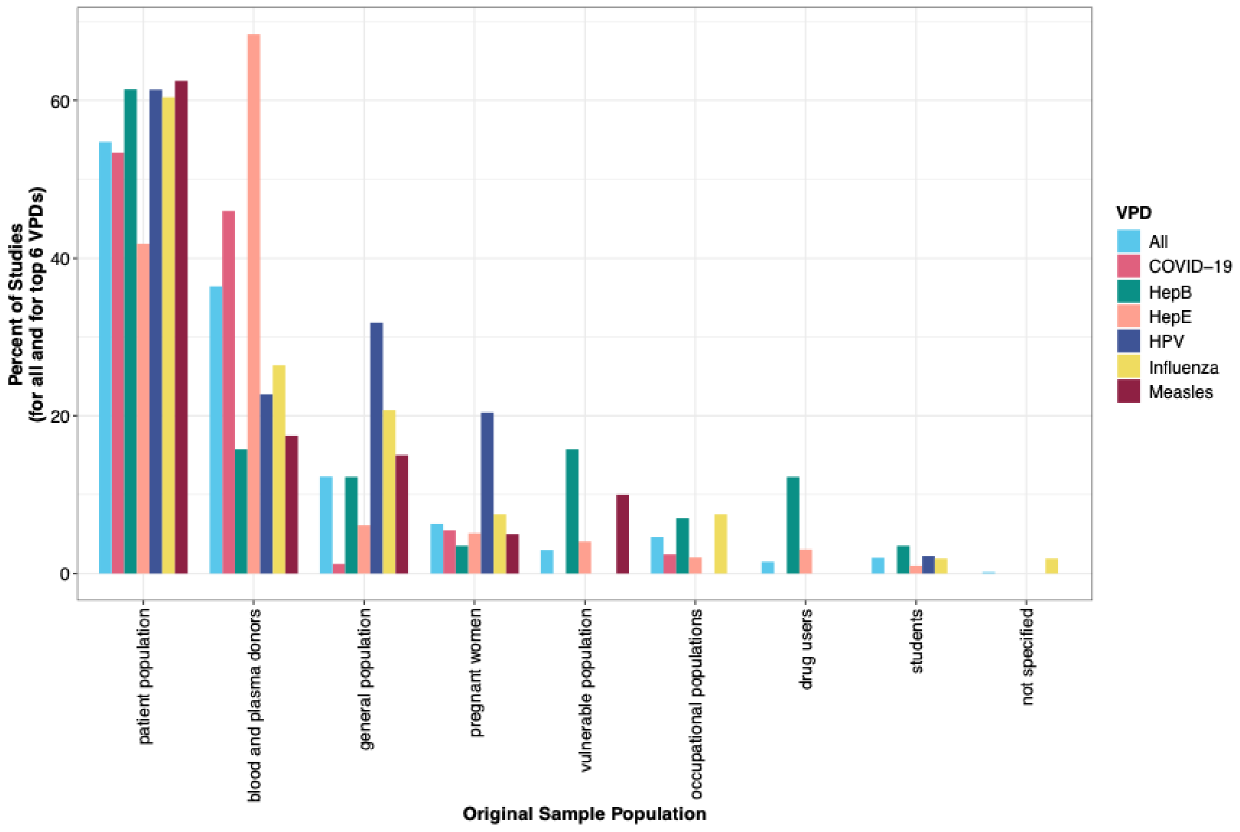

**Figure S2. Time series of studies by publication year based on A) original source population specified in the studies and B) original use for specimens specified in the studies (N = 601 studies).**

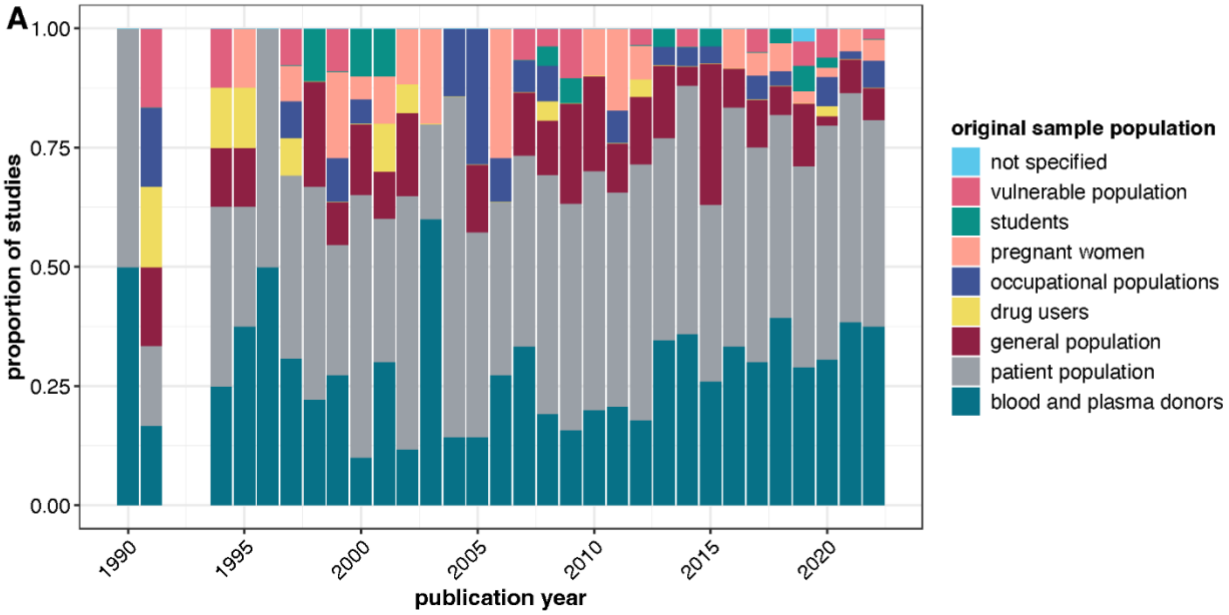

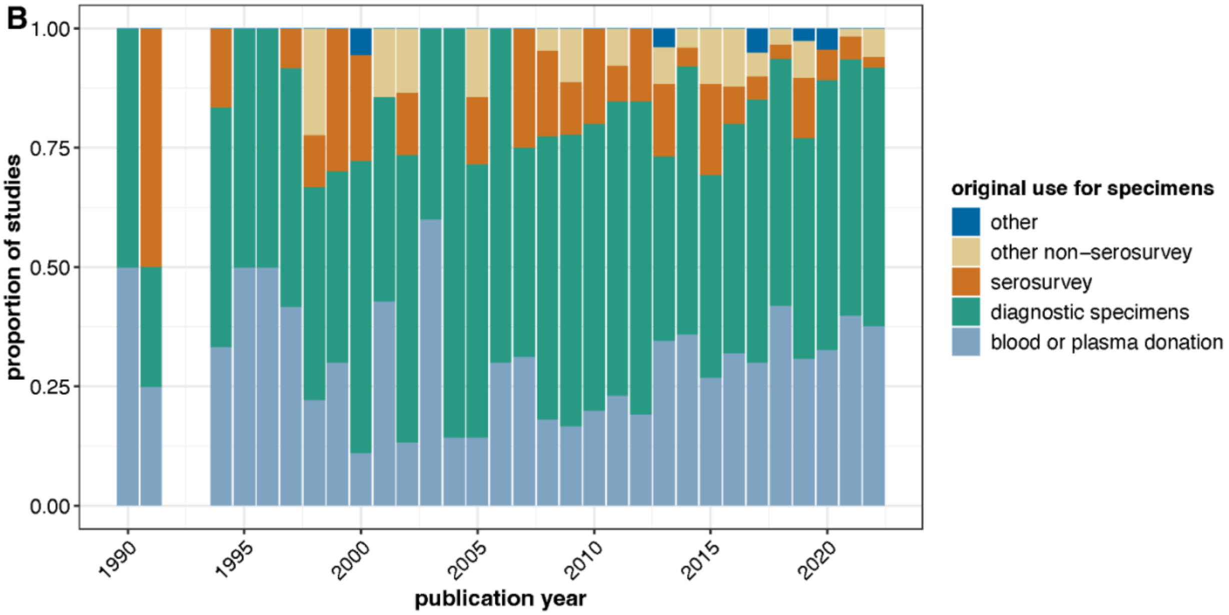

**Figure S3. Histogram of difference between publication year and year of original sample collection.** A) All studies (N = 568 studies), note not all studies listed the year of original sample collection. B) By six most studies VPDs. The red line and number represent the mean number of years.

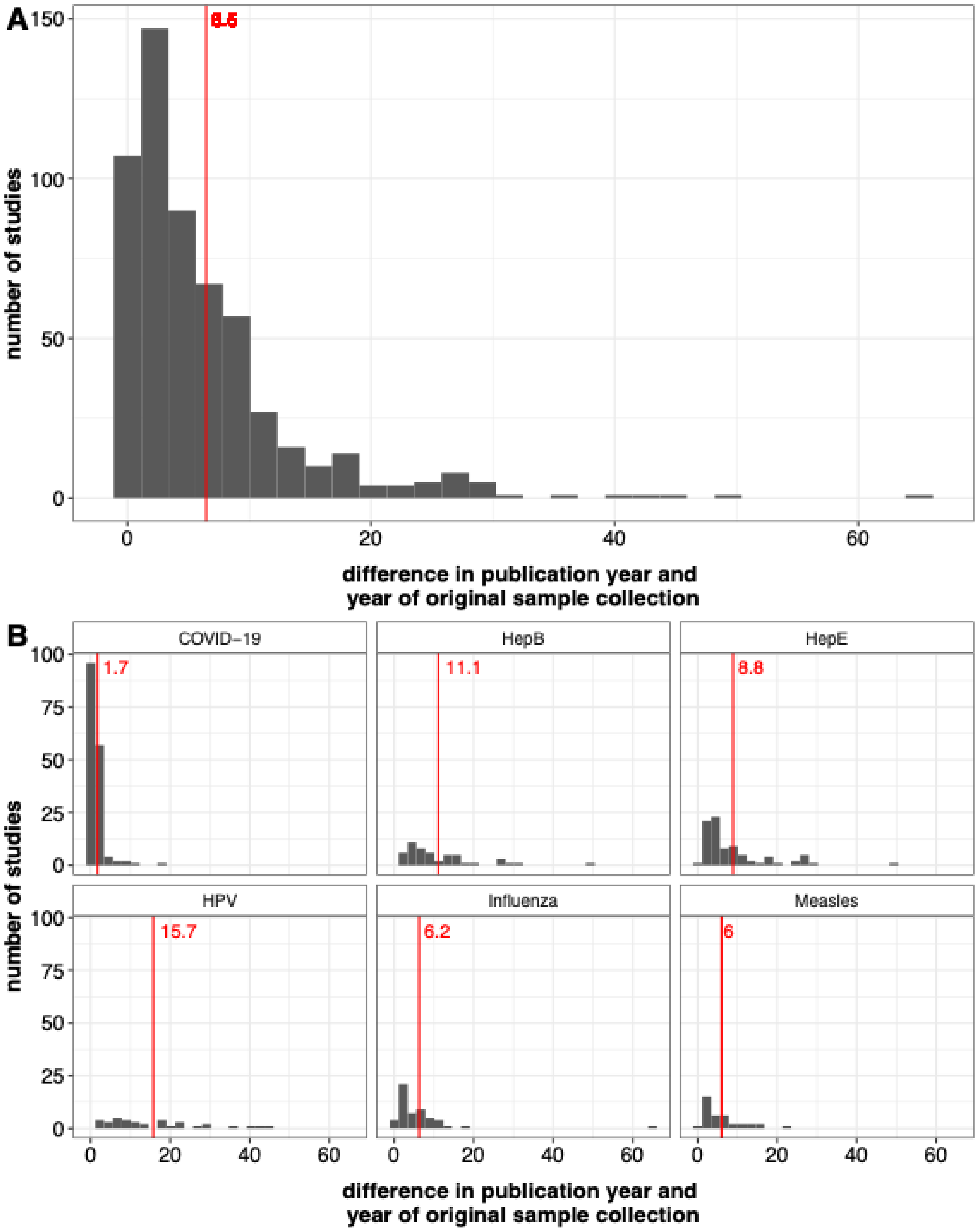

**Table S1. Examples of original sample populations**

| **Original population** | **Examples** |
| --- | --- |
| Patient populations | Definitions: Specimens from routine or specialized laboratory testing from health facilities or diagnostic laboratories;<br>Examples: included clinical subpopulations such as cancer patients or patients on dialysis, patients seeking emergency care, patients attending a specialty health clinic (e.g., HIV clinic), patients with the VPD of interest (e.g., SARS-CoV-2).<br>*Note: Patient populations often used as convenience samples. If it was unclear if the specimen was residual then the article was excluded at the full-text screening stage.* |
| Blood and plasma donors | Definition: Specimens collected from blood or plasma donors as part of routine screening.<br>*Notes:*<br>*Blood and plasma donors were often used as convenience samples. If it was unclear if the specimen was residual then the article was excluded at the full-text screening stage.*<br>Excluded testing of blood or plasma donors for VPDs as part of routine screening within that country; policies for routine screening varied by country and timepoint.<br>Excluded studies where blood or plasma donors were only used as a control group to compare with a non-residual specimen source.<br>Excluded convalescent plasma donors for SARS-CoV-2. |
| Pregnant women | Definition: Specimens collected from pregnant women.<br>Examples: Residual specimens from routine antenatal care testing or cord blood collected at delivery, and pregnant women who were part of a cohort or prior study.<br>Note:<br>*Pregnant women patients often used as convenience samples. If it was unclear if the specimen was residual then the article was excluded at the full-text screening stage.*<br>Excluded routine testing for VPDs (e.g., rubella). |
| Drug users (injection and non-injection) | Definitions: Specimens collected from drug users (injection and non-injection). |

| | |
|---|---|
| | Examples: Residual specimens from drug treatment centers or health clinics, and specimens from drug users collected as part of a cohort or prior study. |
| Students | Definition: Specimens collected from students in a school setting (pre-primary, primary, secondary or university).<br>Examples: Residual specimens collected from a prior study (e.g., vaccination coverage or anemia studies), collected as part of a vaccination program (e.g., prevaccination rubella testing) or collected during a medical check-up of school children. |
| Vulnerable populations | Vulnerable populations: Specimens collected from indigenous populations, migrants, incarcerated individuals, MSM, homeless individuals.<br>Examples: Residual specimens collected during health examinations for asylum seekers or migrants, intake screening of incarcerated individuals, residual specimens from a prior study, or residual specimens collected at a health clinic. |
| Occupational populations | Definitions: Specimens collected from specified occupational populations.<br>Examples: Residual specimens from pre-employment medical assessments (e.g., from healthcare workers, military) and residual specimens collected from occupational populations during a prior study (e.g., pig farmers, fish mongers, tattoo artists). |
| General population | Definition: Specimens collected from populations representative of a country, region or other geographic unit, or not otherwise specified above.<br>Examples: Residual specimens from prior studies (e.g., population-based HIV seroprevalence study).<br>Other research studies such as cohort studies or vaccine clinical trials where the population does not fit into one of the other categories. |

Note: Population categories were not mutually exclusive.

**Table S2. Handling of paper objectives based on article's abstract**

| Original population | Description |
|---|---|
| Describe population seropositivity | Most articles fell into this category, either alone or in combination with other objectives. If the paper was descriptive of seroprevalence, or if no other objectives were clear, or very limited information in the abstract, then this objective was selected alone. |
| Identify risk factors of seropositivity | Abstracts included associations with characteristics of interest (e.g., p-value or odds ratios reported, mention of significant or non-significant differences).<br>If stratified seroprevalence was described in the abstract without an indication the authors conducted a test or were evaluating an association then this objective was not selected. |
| Estimate parameters related to the magnitude or timing of infection or understand transmission dynamics | Abstracts included incidence rates, force of infection, prevalence of recent infection (e.g., pertussis, dengue) and other transmission dynamics estimated using IgG alone or in combination with IgM.<br>If estimates were based only on IgM results and studies where the infection rates described were based on surveillance data then this objective was not selected. |
| Evaluate changes in seropositivity over time | Specimens from multiple timepoints tested as part of the study where at least one time point included residual specimens.<br>If residual specimens from a single time point were qualitatively compared with seroprevalence findings from other studies then this objective was not selected. |
| Describe seroprevalence among a clinical subpopulation | Question of interest was focused on the clinical subpopulation (e.g., seroprevalence among patients with renal cancer or people living with HIV).<br>Pregnant women were not considered clinical subpopulations. If the clinical subpopulation was one of many populations with no direct interest in the findings for that clinical subpopulation then this objective was not selected. |

| | |
|---|---|
| Evaluate health or disease outcomes associated with seropositivity | Association between seropositivity and development of outcomes such as cancer, liver disease, HIV, and severity of the VPD of interest. |
| Evaluate the need for blood donor screening | Studies to evaluate if screening for a VPD should be conducted in settings where it was not already routine practice. |
| Evaluate antibody cross-reactivity | Cross-reactivity of virus variants (e.g., influenza or SARS-CoV-2) or between viruses (e.g., West Nile, yellow fever, Japanese encephalitis). |
| Describe antibody kinetics following vaccination | Given known vaccination status of individuals, e.g., recorded evidence or individuals who were part of a prior study that administered the vaccine, the study evaluated quantitative antibody concentrations following vaccination.<br>Studies comparing infection-induced and vaccine-induced antibody responses fell into both categories (antibody kinetics following vaccination and antibody kinetics following infection). |
| Describe antibody kinetics following infection | Given known infection status of individuals, the study evaluated quantitative antibody concentrations following infection.<br>Studies comparing infection-induced and vaccine-induced antibody responses fell into both categories (antibody kinetics following vaccination and antibody kinetics following infection). |
| Compare population seropositivity between countries or international regions | Smallest administrative unit included for this objective was at the country-level.<br>If comparisons were between regions, districts, states, etc. within the country then this objective was not selected. |
| Compare the use of residual samples to population-based samples | Authors tested specimens from both residual and population-based sources and reported on the comparison.<br>If residual specimens were qualitatively compared with seroprevalence findings from separate population-based study then this objective was not selected. |

| | |
|---|---|
| Describe viral dynamics | Investigating how a virus changes over time (viral strains or lineages). Most relevant for influenza and SARS-Cov-2.<br>If the study estimated serotype/strain/variant at a given time point (cross-sectional study) then this objective was not selected. |

Note 1: Objective of residual specimen serological testing categories are not mutually exclusive.
Note 2: Objective for the residual specimens only as specified in the article's abstract. Objectives for any non-residual specimens that were mentioned in the article's abstract were not captured in data abstraction.
Note 3: We ignored any policy-specific objectives, such as evaluating effectiveness of a mass vaccination program, and abstracted any other objectives in the paper or treated them as 'describe general seropositivity'.

**Table S3. Handling of selection bias in papers using residual specimens**

| Stage | Handling of bias | Description |
|---|---|---|
| Interpretation | Compared to other published estimates | Qualitatively compared seroprevalence findings from the residual specimens with that from other published estimates or administrative sources, typically done in the Discussion section. |
| Analysis | Conducted stratified analysis | Stratified seroprevalence estimates by characteristics associated with seropositivity because if the distribution of the characteristics are not representative of the population of interest then the overall estimate of seroprevalence may be biased (e.g., age, geographic area). |
| Design | Conducted stratified subsampling | Purposefully over or undersampled from specified subgroups to obtain a study population more representative of the target population (e.g., limit the number of specimens from a specific medical ward at a facility) |
| Design | Used inclusion/exclusion criteria for residual samples | Restricted selection of residual specimens by certain characteristics that may influence seroprevalence to obtain a study population more representative of the target population (e.g., exclude specimens from a specific medical ward at a facility, or excluded specimens admitted for respiratory illness). |
| Analysis | Weighted or standardized final results | Seroprevalence estimates are weighted or standardized to align with the distribution of characteristics in the target |

| | | |
|---|---|---|
| | | population (e.g., age, gender, geographic distribution). |
| Analysis | Quantitatively compared to alternative data source | Quantitatively compared seroprevalence findings from the residual specimens with that from other seroprevalence estimates, typically done in the Results section. Some instances may have included formal statistical comparative tests. |
| Analysis | Conducted sensitivity analysis | Reanalysis varying how the input data were handled or other assumptions or parameters used in models.<br>Sensitivity analysis varying thresholds or how equivocal specimens were treated (e.g., treating as positive in primary analysis and negative in sensitivity) was not considered a method of handling of bias. |
| N/A | No selection bias because the residual sample represented the population of interest | Seroprevalence estimated from residual specimens originating from the population of interest (e.g., Hepatitis E serosurvey from residual blood donor specimens to evaluate the need to screen blood donors for Hepatitis E). |

Note: Handling of selection bias categories were not mutually exclusive.

**Table S4**. **Studies selected for data extraction and analysis (N=601 articles)**

| **VPD** | **References** |
|---|---|
| Dengue | [1-31] |
| Diphtheria | [32-50] |
| Hepatitis A (HAV) | [45, 51-88] |
| Hepatitis B (HBV) | [5, 44, 45, 49, 53, 58, 62, 77, 79, 82, 89-135] |
| Hepatitis E (HEV) | [53, 59, 60, 62, 69, 77, 80, 82-84, 114, 136-222] |
| Hemophilus influenza type B (Hib) | [223-225] |
| Human papillomavirus (HPV) | [226-269] |
| Influenza | [270-322] |
| Japanese Encephalitis | [3, 14, 323, 324] |
| Measles | [38, 44, 45, 49, 56, 108, 325-358] |
| Mumps | [38, 45, 108, 327, 331, 347, 352, 355, 357, 359-366] |
| Meningitis | [367, 368] |
| Pertussis | [39, 369-384] |
| Pneumococcal | |
| Poliomyelitis | [39, 385-391] |
| Rotavirus | [392] |
| Rubella | [38, 44, 45, 49, 108, 327, 329, 331, 332, 339, 342, 343, 352, 355-357, 393-409] |
| Sars-CoV-2 | [410-572] |
| Tetanus | [33, 34, 38, 41, 44, 50, 368] |
| Tick-borne Encephalitis | [573, 574] |
| Tuberculosis | |
| Typhoid | [575] |
| Varicella/Herpes Zoster | [44, 45, 49, 61, 108, 329, 576-599] |
| Yellow Fever | [14, 600, 601] |

409. Su, Q., et al., *Assessing the burden of congenital rubella syndrome in China and evaluating mitigation strategies: a metapopulation modelling study.* Lancet Infect Dis, 2021. **21**(7): p. 1004-1013.
410. Adetifa, I.M.O., et al., *Temporal trends of SARS-CoV-2 seroprevalence during the first wave of the COVID-19 epidemic in Kenya.* Nat Commun, 2021. **12**(1): p. 3966.
411. Alandijany, T.A., et al., *Lack of Antibodies to SARS-CoV-2 among Blood Donors during COVID-19 Lockdown: A Study from Saudi Arabia.* Healthcare (Basel), 2021. **9**(1).
412. Alenazi, M.W., et al., *Seroprevalence of COVID-19 in Riyadh city during the early increase of COVID-19 infections in Saudi Arabia, June 2020.* Saudi J Biol Sci, 2022. **29**(6): p. 103282.
413. Alharbi, N.K., et al., *Nationwide Seroprevalence of SARS-CoV-2 in Saudi Arabia.* J Infect Public Health, 2021. **14**(7): p. 832-838.
414. Alzabeedi, K.H., et al., *High Seroprevalence of SARS-CoV-2 IgG and RNA among Asymptomatic Blood Donors in Makkah Region, Saudi Arabia.* Vaccines (Basel), 2022. **10**(8).
415. Amirthalingam, G., et al., *Seroprevalence of SARS-CoV-2 among Blood Donors and Changes after Introduction of Public Health and Social Measures, London, UK.* Emerg Infect Dis, 2021. **27**(7): p. 1795-1801.
416. Amorim Filho, L., et al., *Seroprevalence of anti-SARS-CoV-2 among blood donors in Rio de Janeiro, Brazil.* Rev Saude Publica, 2020. **54**: p. 69.
417. Anderson, E.M., et al., *Seasonal human coronavirus antibodies are boosted upon SARS-CoV-2 infection but not associated with protection.* Cell, 2021. **184**(7): p. 1858-1864 e10.
418. Arabkhazaeli, A., et al., *Positive anti-SARS-CoV-2 rapid serological test results among asymptomatic blood donors.* Transfus Clin Biol, 2022. **29**(1): p. 24-30.
419. Assefa, N., et al., *Seroprevalence of anti-SARS-CoV-2 antibodies in women attending antenatal care in eastern Ethiopia: a facility-based surveillance.* BMJ Open, 2021. **11**(11): p. e055834.
420. Bajema, K.L., et al., *Comparison of Estimated Severe Acute Respiratory Syndrome Coronavirus 2 Seroprevalence Through Commercial Laboratory Residual Sera Testing and a Community Survey.* Clin Infect Dis, 2021. **73**(9): p. e3120-e3123.
421. Bajema, K.L., et al., *Estimated SARS-CoV-2 Seroprevalence in the US as of September 2020.* JAMA Intern Med, 2021. **181**(4): p. 450-460.
422. Bakkour, S., et al., *Minipool testing for SARS-CoV-2 RNA in United States blood donors.* Transfusion, 2021. **61**(8): p. 2384-2391.
423. Basavaraju, S.V., et al., *Serologic Testing of US Blood Donations to Identify Severe Acute Respiratory Syndrome Coronavirus 2 (SARS-CoV-2)-Reactive Antibodies: December 2019-January 2020.* Clin Infect Dis, 2021. **72**(12): p. e1004-e1009.
424. Bingham, J., et al., *Estimates of prevalence of anti-SARS-CoV-2 antibodies among blood donors in South Africa in March 2022.* Res Sq, 2022: p. rs.3.rs–1687679-rs.3.rs–1687679.
425. Boehme, K.W., et al., *Pediatric SARS-CoV-2 Seroprevalence in Arkansas Over the First Year of the COVID-19 Pandemic.* J Pediatric Infect Dis Soc, 2022. **11**(6): p. 248-256.
426. Bogogiannidou, Z., et al., *Repeated Leftover Serosurvey of SARS-CoV-2 IgG Antibodies in Greece, May to August 2020.* Vaccines (Basel), 2021. **9**(5): p. 504-504.
427. Bogogiannidou, Z., et al., *Repeated leftover serosurvey of SARS-CoV-2 IgG antibodies, Greece, March and April 2020.* Euro Surveill, 2020. **25**(31).
428. Bolotin, S., et al., *Assessment of population infection with SARS-CoV-2 in Ontario, Canada, March to June 2020.* Euro Surveill, 2021. **26**(50).
429. Bolotin, S., et al., *SARS-CoV-2 Seroprevalence Survey Estimates Are Affected by Anti-Nucleocapsid Antibody Decline.* J Infect Dis, 2021. **223**(8): p. 1334-1338.
430. Borrega, R., et al., *Cross-Reactive Antibodies to SARS-CoV-2 and MERS-CoV in Pre-COVID-19 Blood Samples from Sierra Leoneans.* Viruses, 2021. **13**(11).
431. Busch, M.P., et al., *Population-Weighted Seroprevalence From Severe Acute Respiratory Syndrome Coronavirus 2 (SARS-CoV-2) Infection, Vaccination, and Hybrid Immunity Among US*

*Blood Donations From January to December 2021.* Clin Infect Dis, 2022. **75**(Suppl 2): p. S254-S263.
432. Buss, L., et al., *Predicting SARS-CoV-2 Variant Spread in a Completely Seropositive Population Using Semi-Quantitative Antibody Measurements in Blood Donors.* Vaccines (Basel), 2022. **10**(9).
433. Buss, L.F., et al., *Three-quarters attack rate of SARS-CoV-2 in the Brazilian Amazon during a largely unmitigated epidemic.* Science, 2021. **371**(6526): p. 288-292.
434. Butler, D., et al., *Confirmed circulation of SARS-CoV-2 in Irish blood donors prior to first national notification of infection.* J Clin Virol, 2022. **146**: p. 105045.
435. Capai, L., et al., *Seroprevalence of SARS-CoV-2 IgG Antibodies in Corsica (France), April and June 2020.* J Clin Med, 2020. **9**(11): p. 3569-3569.
436. Capai, L., et al., *Impact of the Second Epidemic Wave of SARS-CoV-2: Increased Exposure of Young People.* Front Public Health, 2021. **9**: p. 715192.
437. Carlton, L.H., et al., *Charting elimination in the pandemic: a SARS-CoV-2 serosurvey of blood donors in New Zealand.* Epidemiol Infect, 2021. **149**: p. e173.
438. Cassaniti, I., et al., *Seroprevalence of SARS-CoV-2 in blood donors from the Lodi Red Zone and adjacent Lodi metropolitan and suburban area.* Clin Microbiol Infect, 2021. **27**(6): p. 914 e1-914 e4.
439. Castro Dopico, X., et al., *Seropositivity in blood donors and pregnant women during the first year of SARS-CoV-2 transmission in Stockholm, Sweden.* J Intern Med, 2021. **290**(3): p. 666-676.
440. Chanchlani, N., et al., *Adalimumab and Infliximab Impair SARS-CoV-2 Antibody Responses: Results from a Therapeutic Drug Monitoring Study in 11 422 Biologic-Treated Patients.* J Crohns Colitis, 2022. **16**(3): p. 389-397.
441. Chang, L., et al., *The prevalence of antibodies to SARS-CoV-2 among blood donors in China.* Nat Commun, 2021. **12**(1): p. 1383.
442. Charlton, C.L., et al., *Pre-Vaccine Positivity of SARS-CoV-2 Antibodies in Alberta, Canada during the First Two Waves of the COVID-19 Pandemic.* Microbiol Spectr, 2021. **9**(1): p. e0029121.
443. Chau, N.V.V., et al., *Absence of SARS-CoV-2 antibodies in pre-pandemic plasma from children and adults in Vietnam.* Int J Infect Dis, 2021. **111**((Chau N.V.V.; Man D.N.H.; Ty D.T.B.) Hospital for Tropical Diseases, Ho Chi Minh City, Viet Nam): p. 127-129.
444. Chaves, D.G., et al., *Pro-inflammatory immune profile mediated by TNF and IFN-gamma and regulated by IL-10 is associated to IgG anti-SARS-CoV-2 in asymptomatic blood donors.* Cytokine, 2022. **154**((Chaves D.G.,; da Silva Malta M.C.F.; Martins M.L.) Fundação Hemominas, Minas Gerais, Belo Horizonte, Brazil): p. 155874.
445. Chaves, D.G., et al., *SARS-CoV-2 IgG Seroprevalence among Blood Donors as a Monitor of the COVID-19 Epidemic, Brazil.* Emerg Infect Dis, 2022. **28**(4): p. 734-742.
446. Clapham, H.E., et al., *Contrasting SARS-CoV-2 epidemics in Singapore: cohort studies in migrant workers and the general population.* Int J Infect Dis, 2022. **115**: p. 72-78.
447. Couture, A., et al., *Severe Acute Respiratory Syndrome Coronavirus 2 Seroprevalence and Reported Coronavirus Disease 2019 Cases in US Children, August 2020-May 2021.* Open Forum Infect Dis, 2022. **9**(3): p. ofac044.
448. Coyle, P.V., et al., *SARS-CoV-2 seroprevalence in the urban population of Qatar: An analysis of antibody testing on a sample of 112,941 individuals.* iScience, 2021. **24**(6): p. 102646.
449. Damjanovic, A., et al., *Utility of Newborn Dried Blood Spots to Ascertain Seroprevalence of SARS-CoV-2 Antibodies Among Individuals Giving Birth in New York State, November 2019 to November 2021.* JAMA Netw Open, 2022. **5**(8): p. e2227995.
450. Davis, G., et al., *Seroprevalence of SARS-CoV-2 infection in Cincinnati Ohio USA from August to December 2020.* PLoS One, 2021. **16**(7): p. e0254667.
451. Delaugerre, C., et al., *Severe Acute Respiratory Syndrome Coronavirus 2 Seroprevalence Among HIV-Negative Participants Using Tenofovir/Emtricitabine-Based Preexposure Prophylaxis in*

*2020: A Substudy of the French National Agency for Research on AIDS and Viral Hepatitis PREVENIR and Inserm SAPRIS-Sero Cohorts.* Open Forum Infect Dis, 2022. **9**(7): p. ofac188.
452. Dethioux, L., et al., *SARS-CoV-2 seroprevalence in children and their family members, July-October 2020, Brussels.* Eur J Pediatr, 2022. **181**(3): p. 1009-1016.
453. Di Stefano, M., et al., *Low Prevalence of Antibodies to SARS-CoV-2 and Undetectable Viral Load in Seropositive Blood Donors from South-Eastern Italy.* Acta Haematol, 2021. **144**(5): p. 580-584.
454. Dickson, E., et al., *Enhanced surveillance of COVID-19 in Scotland: population-based seroprevalence surveillance for SARS-CoV-2 during the first wave of the epidemic.* Public Health, 2021. **190**: p. 132-134.
455. Dietrich, M.L., et al., *SARS-CoV-2 seroprevalence rates of children seeking medical care in Louisiana during the state stay at home order.* J Clin Virol Plus, 2021. **1**(4): p. 100047.
456. Dingens, A.S., et al., *Serological identification of SARS-CoV-2 infections among children visiting a hospital during the initial Seattle outbreak.* Nat Commun, 2020. **11**(1): p. 4378.
457. Dodd, R.Y., et al., *Characteristics of US Blood Donors Testing Reactive for Antibodies to SARS-CoV-2 Prior to the Availability of Authorized Vaccines.* Transfus Med Rev, 2021. **35**(3): p. 1-7.
458. Drews, S.J., et al., *Resistance of SARS-CoV-2 beta and gamma variants to plasma collected from Canadian blood donors during the spring of 2020.* Transfusion, 2022. **62**(1): p. 37-43.
459. Drews, S.J., et al., *SARS-CoV-2 Virus-Like Particle Neutralizing Capacity in Blood Donors Depends on Serological Profile and Donor-Declared SARS-CoV-2 Vaccination History.* Microbiol Spectr, 2022. **10**(1): p. e0226221.
460. Eldesoukey, N., et al., *SARS-CoV-2 antibody seroprevalence rates among Egyptian blood donors around the third wave: Cross-sectional study.* Health Sci Rep, 2022. **5**(3): p. e634.
461. Erikstrup, C., et al., *Seroprevalence and infection fatality rate of the SARS-CoV-2 Omicron variant in Denmark: A nationwide serosurveillance study.* Lancet Reg Health Eur, 2022. **21**: p. 100479.
462. Eskild, A., et al., *Prevalence of antibodies against SARS-CoV-2 among pregnant women in Norway during the period December 2019 through December 2020.* Epidemiol Infect, 2022. **150**: p. e28.
463. Fischer, B., C. Knabbe, and T. Vollmer, *SARS-CoV-2 IgG seroprevalence in blood donors located in three different federal states, Germany, March to June 2020.* Euro Surveill, 2020. **25**(28).
464. Fischer, B., T. Vollmer, and C. Knabbe, *SARS-CoV-2 IgG seroprevalence in blood donors located in three different federal states, Germany, July 2020 to June 2021 – a follow-up.* medRxiv, 2022((Fischer B.,; Vollmer T.; Knabbe C.) Herz- und Diabeteszentrum NRW, Institut für Laboratoriums- und Transfusionsmedizin, Bad Oeynhausen, Germany).
465. Freeman, M.C., et al., *Immunocompromised Seroprevalence and Course of Illness of SARS-CoV-2 in One Pediatric Quaternary Care Center.* J Pediatric Infect Dis Soc, 2021. **10**(4): p. 426-431.
466. Furukawa, K., et al., *Seroepidemiological Survey of the Antibody for Severe Acute Respiratory Syndrome Coronavirus 2 with Neutralizing Activity at Hospitals: A Cross-sectional Study in Hyogo Prefecture, Japan.* JMA J, 2021. **4**(1): p. 41-49.
467. Galipeau, Y., et al., *Relative Ratios of Human Seasonal Coronavirus Antibodies Predict the Efficiency of Cross-Neutralization of SARS-CoV-2 Spike Binding to ACE2.* EBioMedicine, 2021. **74**: p. 103700.
468. Gallian, P., et al., *Lower prevalence of antibodies neutralizing SARS-CoV-2 in group O French blood donors.* Antiviral Res, 2020. **181**: p. 104880.
469. George, J.A., et al., *Sentinel seroprevalence of SARS-CoV-2 in Gauteng Province, South Africa, August - October 2020.* S Afr Med J, 2021. **111**(11): p. 1078-1083.

**Table S5. Paper objective excluding articles describing population seropositivity (N = 117 studies).** Note the categories below are not mutually exclusive meaning that one study could have conducted testing for multiple pathogens and have had multiple objectives.

| Objective | Total, N (column %) |
|---|---|
| Describe seroprevalence among a clinical subpopulation | 69 (59.0) |
| Identify risk factors of seropositivity | 38 (32.5) |
| Evaluate outcomes of seropositivity | 19 (16.2) |
| Evaluate the need for blood donor screening | 16 (13.7) |
| Estimate parameters related to the magnitude or timing of infection or understand transmission dynamics | 16 (13.7) |
| Describe antibody kinetics following infection | 12 (10.3) |
| Describe antibody kinetics following vaccination | 11 (9.4) |
| Evaluate changes in seropositivity over time | 10 (8.5) |
| Evaluate antibody cross-reactivity | 10 (8.5) |
| Compare population seropositivity between countries or international regions | 3 (2.6) |
| Describe viral dynamics | 2 (1.7) |
| Compare the use of residual samples to population-based samples | 1 (0.9) |
| Total | 117 (100) |

**Table S6. Collection site and Storage sites of the sample (N = 601 studies).** Note, the categories below are not mutually exclusive meaning that one article could have multiple collection or storage sites.

| | **No. of papers (%)** |
|---|---|
| **Collection Site of Specimens** | |
| Blood and plasma donation site | 208 (34.6) |
| Hospital/ Clinic | 207 (34.4) |
| Diagnostic center/ Laboratory | 163 (27.1) |
| Household | 33 (5.5) |
| Unknown | 79 (13.1) |
| Other [a] | 12 (2.0) |
| **Storage Site** | |
| Diagnostic center/laboratory | 235 (39.1) |
| Research biorepository | 199 (33.1) |
| Blood/plasma donation center | 192 (32.0) |
| Unspecified biorepository | 30 (5.0) |

a. Other collection sites included homeless shelters, schools and prisons.

**Table S7. Number of articles utilizing residual specimens from serological surveillance systems**

| Identified Serological Surveillance System | Number of articles |
|---|---|
| Health Protection Agency serological surveillance programme (England) | 12 |
| European Sero-Epidemiology Network (ESEN) | 11 |
| National Centre for Immunisation Research and Surveillance (NCIRS) (Australia) | 4 |
| Vietnam serosurveillance (serum biobank project) | 1 |

**Table S8. Meta-data linked to residual samples by the original use of the sample (N = 601 studies).** Note, the categories below are not mutually exclusive meaning that one article could have residual samples linked to multiple categories of meta-data.

| **Meta-data linked to residual samples** | **Total n (column %)** | **Original use of specimen n (column %)** | | | |
|---|---|---|---|---|---|
| | | **Blood or plasma donation** | **Diagnostic specimens** | **Serosurvey** | **Other non-serological survey** |
| Basic demographic data (i.e., age, sex, race, ethnicity) | 567 (94.3) | 194 (88.2) | 356 (96.0) | 62 (95.4) | 32 (100) |
| Extended demographic data (e.g., residence, occupation, SES) | 284 (47.3) | 102 (46.4) | 158 (42.6) | 36 (55.4) | 15 (46.9) |
| Epidemiologic data on VPD (e.g., vaccination status or infection history) | 106 (17.6) | 25 (11.4) | 66 (17.8) | 21 (32.3) | 9 (28.1) |
| Epidemiologic data not on VPD (e.g., vaccination status or infection history) | 97 (16.1) | 33 (15) | 62 (16.7) | 11 (16.9) | 7 (21.9) |
| None | 25 (4.2) | 19 (8.6) | 9 (2.4) | 3 (4.6) | 0 (0) |
| Total | 601 (100) | 220 (100) | 371 (100) | 65 (100) | 32 (100) |

**Table S9. Permissions and ethical considerations by original use of specimens (N=601 articles).** Note, the categories below are not mutually exclusive meaning that one article could have discussed multiple permission or ethical considerations.

| Permission and Ethical Considerations | Total n (column %) | Original use of specimen n (column %) | | | |
|---|---|---|---|---|---|
| | | Blood or plasma donation | Diagnostic specimens | Serosurvey | Other non-serological survey |
| Approved by ethics board/committee | 423 (70.4) | 149 (67.7) | 257 (69.3) | 43 (66.2) | 29 (90.6) |
| Broad consent at time of specimen collection | 199 (33.1) | 91 (41.4) | 94 (25.3) | 27 (41.5) | 16 (50.0) |
| Samples de-identified | 199 (33.1) | 60 (27.3) | 137 (36.9) | 13 (20.0) | 10 (31.3) |
| No statement about permissions or ethics | 88 (14.6) | 37 (16.8) | 58 (15.6) | 14 (21.5) | 2 (6.3) |
| Waiver on re-consenting | 45 (7.5) | 14 (6.4) | 29 (7.8) | 2 (3.1) | 0 (0) |
| Exempt as public health surveillance | 20 (3.3) | 4 (1.8) | 15 (4.0) | 2 (3.1) | 0 (0) |
| Other | 8 (1.3) | 3 (1.4) | 4 (1.1) | 0 (0) | 0 (0) |
| Reconsent given | 2 (0.3) | 0 (0) | 1 (0.3) | 1 (1.5) | 0 (0) |
| Total | 601 (100) | 220 (100) | 371 (100) | 65 (100) | 32 (100) |
